# The prognostic value of blood-based p-tau217 levels on progression to clinical impairment

**DOI:** 10.64898/2026.02.06.26345770

**Authors:** Rachel F. Buckley, Diana L. Townsend, Colin J. Birkenbihl, Madison Cuppels, Gillian T. Coughlan, Mabel T. Seto, Jane A. Brown, Michael J. Properzi, Merle C. Hönig, Annie Li, Aaron P. Schultz, Jasmeer Chhatwal, Hyun-Sik Yang, Steven Arnold, Pia Kivisäkk, Bryan D. James, Sid O’Bryant, Robert A. Rissman, Melissa Petersen, Jessica Z. K. Caldwell, Tobey Betthauser, Julie Elisabeth Oomens, Maria Carrigan, Brian Healy, Jorge Garcia Condado, Sterling Johnson, Wai-Ying Wendy Yau, Oliver Langford, Michelle Farrell, Rebecca E. Amariglio, Dorene M. Rentz, Kathryn V. Papp, Timothy J. Hohman, Michael Donohue, Paul S. Aisen, Keith A. Johnson, Reisa A. Sperling

**Affiliations:** Department of Neurology, Mass General Brigham, Boston, MA, USA; Melbourne School of Psychological Sciences, University of Melbourne, Melbourne, Australia; Research Center Juelich, Institute for Neuroscience and Medicine II, Juelich, Germany; Rush Alzheimer’s Disease Center, Rush University Medical Center, Chicago, IL, USA; University of North Texas Health Science Center, Denton, TX, USA; University of Southern California, Los Angeles, CA, USA; University of North Texas Health Science Center, Fort Worth, TX, USA; University of Wisconsin–Madison, Madison, WI, USA; Department of Neurology, Amsterdam UMC, Amsterdam, Netherlands; Vanderbilt University Medical Center, Nashville, TN, USA; Department of Radiology, Mass General Brigham, Boston, MA, USA

## Abstract

Plasma p-tau217 closely tracks amyloid-β (Aβ) pathology, yet its ability to predict long-term clinical progression in cognitively unimpaired (CU) adults remains uncertain. We analyzed harmonized data from 2,705 CU participants (Age_mean_=69.8±7years; Female=63%) across six longitudinal cohorts with up to 13.5 years of follow-up. Cox models evaluated associations between p-tau217 and progression to a clinical diagnosis of cognitive impairment, while natural cubic spline models assessed associations with longitudinal decline on a cognitive composite. Higher p-tau217 was associated with increased risk of progression (hazard-ratio[HR]=1.38; 95%CI:1.31–1.44), independent of demographics and *APOE*ε4, and in models with Aβ-PET (HR=1.30; 95%CI:1.23–1.38). Very high p-tau217 levels (>2.5SD) were associated with 61%[95%CI:53-68%] absolute risk of progression over 10 years. Elevated p-tau217 associated with accelerated cognitive decline, both independent of, and synergistic with, greater Aβ-PET. These findings establish plasma p-tau217 as a robust prognostic marker in preclinical AD and support its value in future individualized risk prediction.

---

Blood-based biomarkers have fundamentally transformed Alzheimer’s disease (AD) research and have become a common feature of observational cohorts and prevention trials^1-5^. With the recent regulatory approval of a plasma phosphorylated tau at threonine 217 (p-tau217) assay for clinical use^6^, we are entering a new era in which plasma p-tau217 is increasingly incorporated into routine clinical care across many medical settings, supporting diagnostic confirmation and treatment decision-making in symptomatic patients^7^. As the possibility of treating AD at earlier preclinical stages is currently being tested in large-scale trials^8, 9^, and as plasma p-tau217 becomes poised for broader adoption in both primary care and specialty settings^10^, a critical question emerges: what does an elevated p-tau217 level mean for a cognitively unimpaired (CU) individual’s absolute risk of developing impairment over clinically relevant time frames from 2 to 10 years? Prior efforts have largely relied on β-amyloid (Aβ) PET imaging or post-mortem neuropathological evidence^11-13^, given the availability of large samples now with PET biomarker or pathologic data available. Early studies combined both data types to model progression across biomarker-defined disease stages and estimate risk of progression to mild cognitive impairment (MCI) or dementia^11^. More recently, a large study using data from the Mayo Clinic Study of Aging (MCSA) provided lifetime and 10-year absolute risk estimates using continuous Aβ-PET Centiloid values in conjunction with demographic factors^13^. The authors demonstrated that increasing Aβ burden robustly predicted progression to cognitive impairment, while age exerted an even stronger modifying effect. These studies represented a major conceptual advance by translating biomarker abnormalities into absolute risk estimates that are directly interpretable for patients and clinicians.

The clinical relevance of 10-year risk estimates to a diagnosis of cognitive impairment is clear; follow-up in both research and clinical settings is inherently constrained, and shorter-term risk horizons offer greater clinical utility than lifetime risk estimates, which are less applicable in real-world decision-making. There is a parallel need to understand shorter-term risk over 2- and 5-year windows. Although prevention trials rarely use diagnostic progression as a primary endpoint, knowledge of absolute short-term risk remains critical for trial enrichment strategies. Specifically, identifying biomarker thresholds that capture individuals at highest imminent risk will improve efficiency, accelerate therapeutic development, and provide critical information for clinicians and patients to inform shared decision-making once disease-modifying therapies are available.

In light of this need, estimating absolute risk of progression using blood-based biomarkers is a burgeoning area of research. A recent large, population-based study demonstrated that serum levels of p-tau217 (alongside serum levels of NfL and GFAP) robustly predicted incident all-cause and AD dementia over 10 years, with strong discrimination and high negative predictive value (NPV), but low positive predictive value (PPV)^14^. This work, however, did not provide time-specific absolute risk estimates based on biomarker level estimates of AD pathology. This gap is increasingly consequential as clinical and research frameworks have shifted decisively toward biomarker-driven diagnoses of AD. As primary care physicians and specialists begin to rely on plasma p-tau217 to inform prognosis in CU older adults, it is essential to define how biomarker levels translate into individualized risk of progression to clinical impairment over clinically meaningful time horizons, including mild cognitive impairment (MCI) or equivalent early functional change^15^. Here, we address this critical gap by quantifying the prognostic utility of plasma p-tau217 in CU older adults over 2-, 5-, and 10-year windows. To contextualize these estimates within established biomarker frameworks, we additionally evaluate risk in relation to Aβ-PET Centiloid values. By providing time-specific absolute risk estimates anchored in both plasma and imaging biomarkers, this work aims to inform clinical decision-making and trial design in preclinical AD.

## Results

We accessed, harmonized and pooled data from 2,705 cognitively unimpaired participants (mean (s.d; range) age, 69.8 (7.0; 46-98.1)□years; 1,704 women (63%); Table 1) from six different cohorts; the Alzheimer’s Disease Neuroimaging Initiative (ADNI), the Anti-Amyloid Treatment in Asymptomatic Alzheimer’s (A4) Study and its observational arm the Longitudinal Evaluation of Amyloid Risk and Neurodegeneration (LEARN) Study, the Harvard Aging Brain Study (HABS), the Health & Aging Brain Study-Health Disparities (HABS-HD), and the Wisconsin Registry for Alzheimer’s Prevention (WRAP). All participants were cognitively unimpaired at baseline, had a baseline plasma p-tau217 value and Aβ-PET Centiloid value available as well as at least one follow-up clinical assessment, which provided clinical diagnostic status (overall follow-up in years mean (s.d; range): 5.1 (2.7; 0.1-13.5) years (see Supplementary Figure A for the cohort-specific and full wave diagram of time-to-event). 139 participants (5%) had at least 10 years of clinical follow-up, with 36 of those progressing to dementia (26%) during their study participation (mean time to event = 7.3 (0.7-12.5) years). We found significantly different events and follow-up by cohort, with significantly fewer events in LEARN, WRAP and HABS-HD and shorter follow-up in HABS-HD.

**Table 1.** Demographic characteristics of the overall sample and by cohort.

| Demographics | Overall<br>N=2,705 | A4<br>N=954 | ADNI<br>N=253 | HABS<br>N=287 | HABS-HD<br>N=548 | LEARN<br>N=509 | WRAP<br>N=154 | p-val |
| --- | --- | --- | --- | --- | --- | --- | --- | --- |
| Age (yrs)<br>Mean (SD)<br>[Range] | 69.8 (7.0)<br>[46.0, 98.1] | 72.1 (4.8)<br>[65.2, 86.2] | 72.9 (7.3)<br>[55.2, 98.1] | 71.4 (7.8)<br>[50.0, 89.0] | 65.11 (7.8)<br>[51.0, 85.4] | 70.6 (4.3)<br>[65.0, 86.6] | 61.6 (6.4)<br>[46.0, 75.0] | <0.001 |
| Female (%) | 63 | 60 | 57 | 61 | 73 | 62 | 68 | <0.001 |
| Education (yrs)<br>Mean (SD)<br>[Range] | 16.0 (3.2)<br>[0.0, 30.0] | 16.5 (2.8)<br>[7.0, 27.0] | 16.7 (2.4)<br>[11.0, 20.0] | 16.0 (2.9)<br>[8.0, 20.0] | 14.0 (4.1)<br>[0.0, 20.0] | 16.8 (2.6)<br>[8.0, 30.0] | 16.4 (2.7)<br>[12.0, 25.0] | <0.001 |
| APOEε4+ (%) | 39 | 58 | 37 | 28 | 28 | 22 | 42 | <0.001 |
| Black/African<br>American n (%) | 249 (9%) | 16 (2%) | 16 (6%) | 42 (15%) | 159 (29%) | 12 (2%) | 4 (3%) | <0.001 |
| White (%) | 92 | 94 | 87 | 82 | 70 | 94 | 96 |  |
| Hispanic n (%) | 234 (9%) | 25 (3%) | 16 (6%) | 4 (1%) | 171 (31%) | 17 (3%) | 1 (1%) | <0.001 |
| Aβ+ >25CL (%) | 43 | 90 | 36 | 24 | 19 | 2.2 | 13 | <0.001 |
| p-tau217 (z-score)<br>Mean (SD)<br>[Range] | 1.04 (1.77)<br>[-3.25,<br>9.72] | 2.16 (1.88)<br>[-2.43, 9.72] | 0.94 (1.63)<br>[-3.25,<br>6.41] | 0.77 (1.45)<br>[-2.08,<br>5.22] | 0.46 (1.29)<br>[-2.44, 6.20] | -0.02 (1.07)<br>[-2.54,<br>6.62] | 0.34 (1.32)<br>[-2.71,<br>4.40] | <0.001 |
| Total follow-up<br>(yrs)<br>Mean (SD)<br>[Range] | 5.12 (2.73)<br>[0.12, 13.5] | 4.86 (2.16)<br>[0.32, 8.65] | 3.59 (2.00)<br>[0.45, 11.4] | 8.00 (3.55)<br>[0.86, 13.5] | 2.61 (1.00)<br>[1.37, 7.00] | 5.56 (2.12)<br>[0.12, 8.91] | 8.07 (2.00)<br>[1.83,<br>11.40] | <0.001 |
| Number of<br>progression<br>events n(%) | 518 (19%) | 259 (27%) | 67 (26%) | 62 (22%) | 64 (12%) | 55 (11%) | 11 (7%) | <0.001 |

**Table 1.** Demographic characteristics of the cohorts and overall sample.

| Demographics | Overall<br>N=2,705 | ADNI<br>N=253 | HABS<br>N=287 | A4<br>N=954 | LEARN<br>N=509 | WRAP<br>N=154 | HABS-HD<br>N=548 |
| --- | --- | --- | --- | --- | --- | --- | --- |
| Age (yrs)<br>Mean (SD)<br>[Range] | 69.8 (7.0)<br>[46.0, 98.1] | 72.9 (7.3)<br>[55.2, 98.1] | 71.4 (7.8)<br>[50.0, 89.0] | 72.1 (4.8)<br>[65.2, 86.2] | 70.6 (4.3)<br>[65.0, 86.6] | 61.6 (6.4)<br>[46.0, 75.0] | 68.1 (7.8)<br>[51.0, 85.4] |
| Female (%) | 63 | 57 | 61 | 60 | 62 | 68 | 73 |
| Education (yrs)<br>Mean (SD)<br>[Range] | 16.0 (3.2)<br>[0.0, 30.0] | 16.7 (2.4)<br>[11.0, 20.0] | 16.0 (2.9)<br>[8.0, 20.0] | 16.5 (2.8)<br>[7.0, 27.0] | 16.8 (2.6)<br>[8.0, 30.0] | 16.4 (2.7)<br>[12.0, 25.0] | 14.0 (4.1)<br>[0.0, 20.0] |
| *APOE*ε4+ (%) | 39 | 37 | 28 | 58 | 22 | 42 | 28 |
| Black/African<br>American n (%) | 249 (9%) | 16 (6%) | 42 (15%) | 16 (2%) | 12 (2%) | 4 (3%) | 159 (29%) |
| White (%) | 92 | 87 | 82 | 94 | 94 | 96 | 98 |
| Hispanic n (%) | 234 (9%) | 16 (6%) | 4 (1%) | 25 (3%) | 17 (3%) | 1 (1%) | 171 (31%) |
| Amyloid+ (%) | 43 | 36 | 24 | 90 | 2.2 | 13 | 19 |
| p-tau217 (z-score)<br>Mean (SD)<br>[Range] | 1.04 (1.77)<br>[-3.25, 9.72] | 0.94 (1.63)<br>[-3.25, 6.41] | 0.77 (1.45)<br>[-2.08, 5.22] | 2.16 (1.88)<br>[-2.43, 9.72] | -0.02 (1.07)<br>[-2.54, 6.62] | 0.34 (1.32)<br>[-2.71, 4.40] | 0.46 (1.29)<br>[-2.44, 6.20] |
| Follow-up (yrs)<br>Mean (SD)<br>[Range] | 4.74 (2.70)<br>[0.12, 13.38] | 3.12 (1.60)<br>[0.45, 9.10] | 7.35 (3.70)<br>[0.76, 13.38] | 4.86 (2.16)<br>[0.32, 8.65] | 5.17 (2.32)<br>[0.12, 8.68] | 7.96 (2.05)<br>[1.83, 11.44] | 2.61 (0.95)<br>[1.37, 7.00] |
| Number of<br>MCI/Dem/2<br>consecutive 0.5<br>CDR events n(%) | 518 (19%) | 67 (26%) | 62 (22%) | 259 (27%) | 55 (11%) | 11 (7%) | 64 (12%) |

### The contribution of p-tau217 to the prediction of progression to cognitive impairment

We ran a series of Cox Proportional Hazards models. Our primary outcome was clinical evidence of cognitive impairment (a within-cohort diagnosis of mild cognitive impairment (MCI), dementia or 2+ consecutive ≥0.5 Global Clinical Dementia Rating (CDR) Scale; see Supplemental Table 1 for breakdown of these categories by cohort). Plasma p-tau217 was modeled as both a continuous and categorical predictor, with four categories (Low, Intermediate, High, and Very High) were defined using a conditional quantiling approach (see Methods for details) that anchored p-tau217 distributions to established Aβ-PET Centiloid (CL) cut points of 10, 25, and 60 CL (see Supplementary Figure B for comparisons between p-tau217 and Aβ-CL). The aim was to ensure that p-tau217 groups corresponded to Aβ-PET-defined thresholds cohorts and assay platforms rather than arbitrary percentile-based groupings. These groups were defined as Low (<−0.5SD; corresponding to <10 AβCL, Intermediate (0.5-1.0SD; °10-24 AβCL), High (1.1-2.4SD; 25-60 AβCL) and Very High: (>+2.5SD; >60 AβCL). The aim was to ensure that p-tau217 groups corresponded to Aβ-PET-defined thresholds cohorts and assay platforms rather than arbitrary percentile-based groupings (see Supplementary Figure B). We chose these CL cut-points given their association with the lowest threshold to show meaningful change (10CL)^16^, a more traditional Aβ+ cut-point (25CL)^17, 18^, and a higher Aβ threshold (60CL) that is typically associated with early neocortical tau^19^. 516 participants were assigned to the Low category, 1,089 were Intermediate, 594 were High and 506 were Very High (sample difference characteristics are found in Supplementary Table 2). Covariates for all models were cohort, age, sex, years of education and *APOE*_ε_4 status.

In the full sample, there were 518 events, with a mean time to event at 3.8 (2.2, 0.4-12.5) years. We found that, of those who progressed, 95% progressed to MCI while only a minority (1.3%) progressed directly to a diagnosis of dementia. An increase of one standard deviation in p-tau217 was associated with a 37% (95% CI: 30-44, *p*<0.0001; Figure 1A) increased risk of progression to cognitive impairment. In the same model, we found older age, male sex and lower education to contribute to risk of progression (see Figure 1B and Supplementary Table 3A for full model estimates). When considering p-tau217 categories, we found the High and Very High groups were greatest contributors to risk of progression, with hazard ratios at 2.15 (95% CI: 1.53-3.03, *p*<0.0001) and 4.12 (95% CI: 2.94, 5.77, *p*<0.0001) relative to the Low p-tau217 group, respectively (see Figures 1C and 1D).

**Figure 1.**
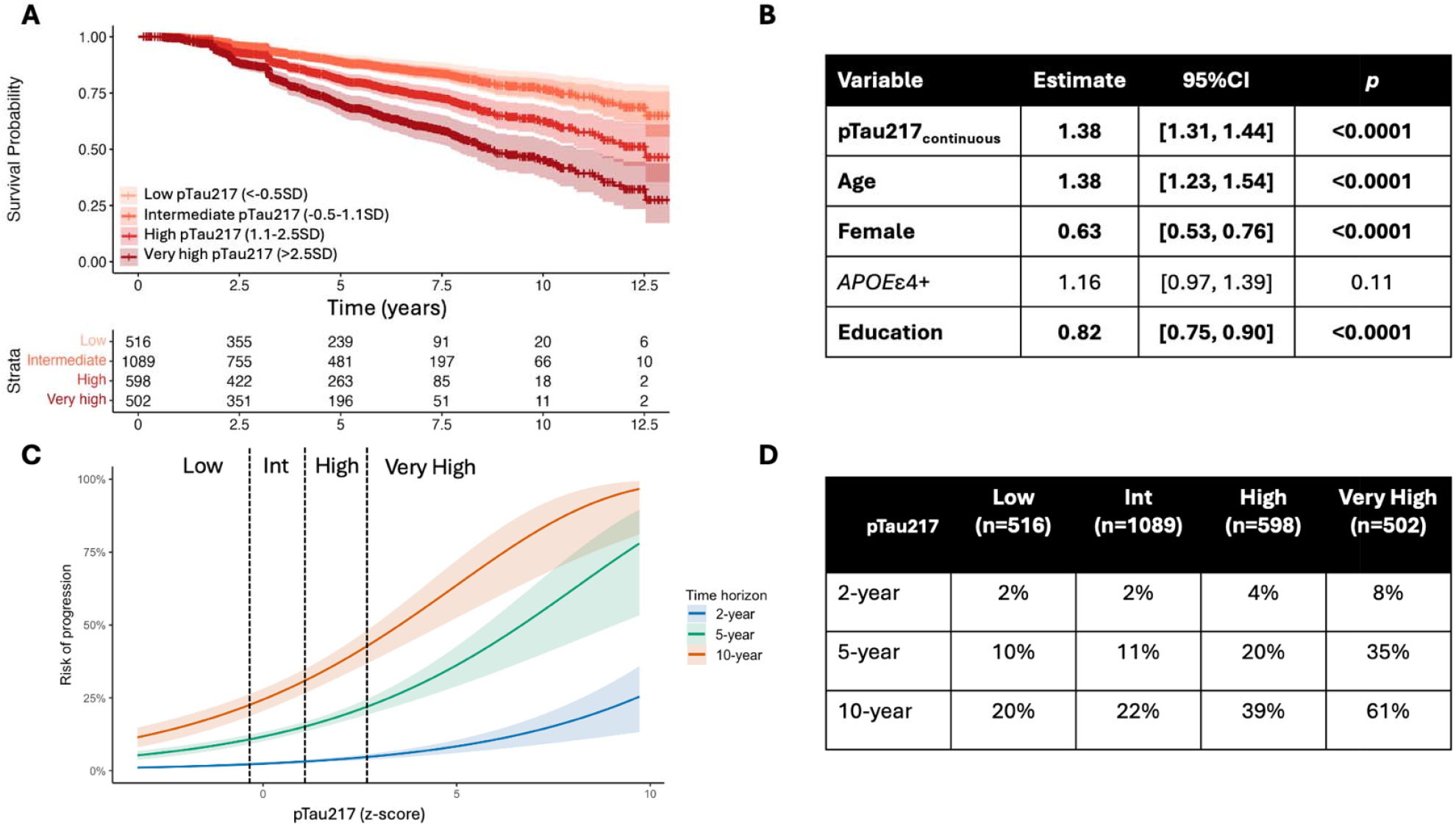
(A) Survival curves for clinical progression to cognitive impairment by p-tau_217_ group (Low [corresponding to estimated <10 AβCL], Intermediate [10-24 AβCL], High [25-60 AβCL] and Very High [>60 AβCL]). (B) Standardized model estimates of the main effects of predictors of interest, including continuous plasma p-tau217, in the survival model (all continuous predictors were standardized [mean = 0, SD = 1]). (C) Model-based 2-, 5-, and 10-year absolute risk by p-tau_217_ group. (D) Tabular absolute risk values by p-tau217 group.

We also estimated absolute risk across the different p-tau217 categories for the three time-horizons (2-, 5-, and 10-years) in a reference group that was the average age, sex, education and *APOE*_ε_4 status of the sample. Two-year absolute risk was found to be very low regardless of the p-tau217 category (2-8%), while 5-year absolute risk was higher across the categories at 10%_Low,_ 11%_Intermediate,_ 20%_High_, and 35%_Very High,_ respectively. For the 10-year time horizon, the following absolute risk was reported at 20%_Low,_ 22%_Intermediate,_ 39%_High_, and 61%_Very High_. Absolute risk was consistently higher in the older age bands (Figure 2A) and in males (Figure 2B). The model showed strong long-term risk discrimination (10-year AUC = 0.88) and good overall predictive performance (C-index = 0.73). Calibration error was modest (Brier score = 0.20), indicating reasonable agreement between predicted and observed risk. Limiting our analyses to only those who progressed to MCI (n=2388, # events=491), we found no major diversion of our estimates from the full model. When comparing Cox Proportional Hazards models stratified by cohort, we found HRs were consistent across all cohorts (HRs: 1.31-1.44) excepting the WRAP cohort that showed a higher HR and a much broader CI 95% (see Supplementary Figure C and cohort-specific analyses in Supplementary Tables 3A-E).

**Figure 2.**
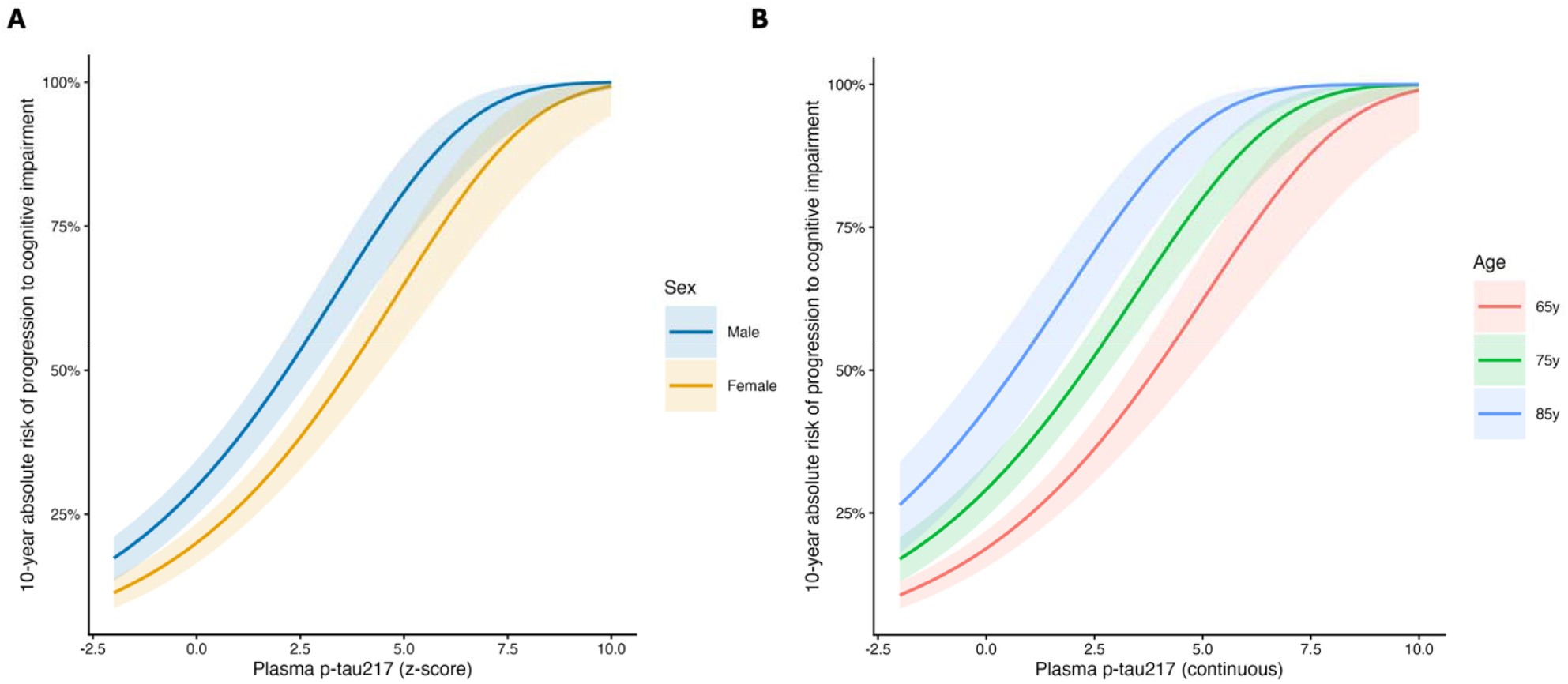
Ten-year absolute risk of progression to cognitive impairment across the plasma p-tau217 spectrum (with 95% confidence interval bands), stratified by (A) sex and (B) age. All estimates are averaging across cohort, *APOE*ε4 status, education, sex and age (*emmeans*).

### The contribution of p-tau217 to the prediction of progression to cognitive impairment in <10 Aβ-CL

In a secondary analysis in only those who were below 10 Aβ-CL on PET at baseline (n=1,124), there were 111 events. We found an increase of one standard deviation in p-tau217 was associated with a 28% (95% CI: 9-50, *p*=0.002; see Figure 3) increased risk of progression to cognitive impairment. In the same model, we found male sex and lower education to contribute to risk of progression (see Supplementary Table 3C for full model estimates). When considering p-tau217 categories, we found only the Very High group was associated with risk of progression, with a hazard ratio of 2.56 (95% CI: 1.11-5.92, *p*=0.03) relative to the Low p-tau217 group. It is important to note that the confidence intervals were much larger, particularly past >2.5SD p-tau217, denoting greater uncertainty in the model from many fewer progression events (n=8) past this range (see Supplementary Table 4 for contingency tables). We also estimated absolute risk across the different p-tau217 categories for the three time-horizons (2-, 5-, and 10-years) in a reference group that was the average age, sex, education and *APOE*_ε_4 status of the sample. Two-year absolute risk was again found to be very low regardless of the p-tau217 category (2-8%), while 5-year absolute risk was higher across the categories at 8%_Low,_ 15%_Intermediate,_ 21%_High_, and 33%_Very High,_ respectively. For the 10-year time horizon, the following absolute risk was reported at 11%_Low,_ 19%_Intermediate,_ 27%_High_, and 41%_Very High_. It is important to note again that the confidence around our estimates of absolute risk became very wide after the 2.5SD p-tau217 level, indicating low confidence in this end of the continuum. While the AUC and C-index were consistent with previous models, the calibration error was higher (Brier score = 0.30), indicating less agreement between predicted and observed risk.

**Figure 3.**
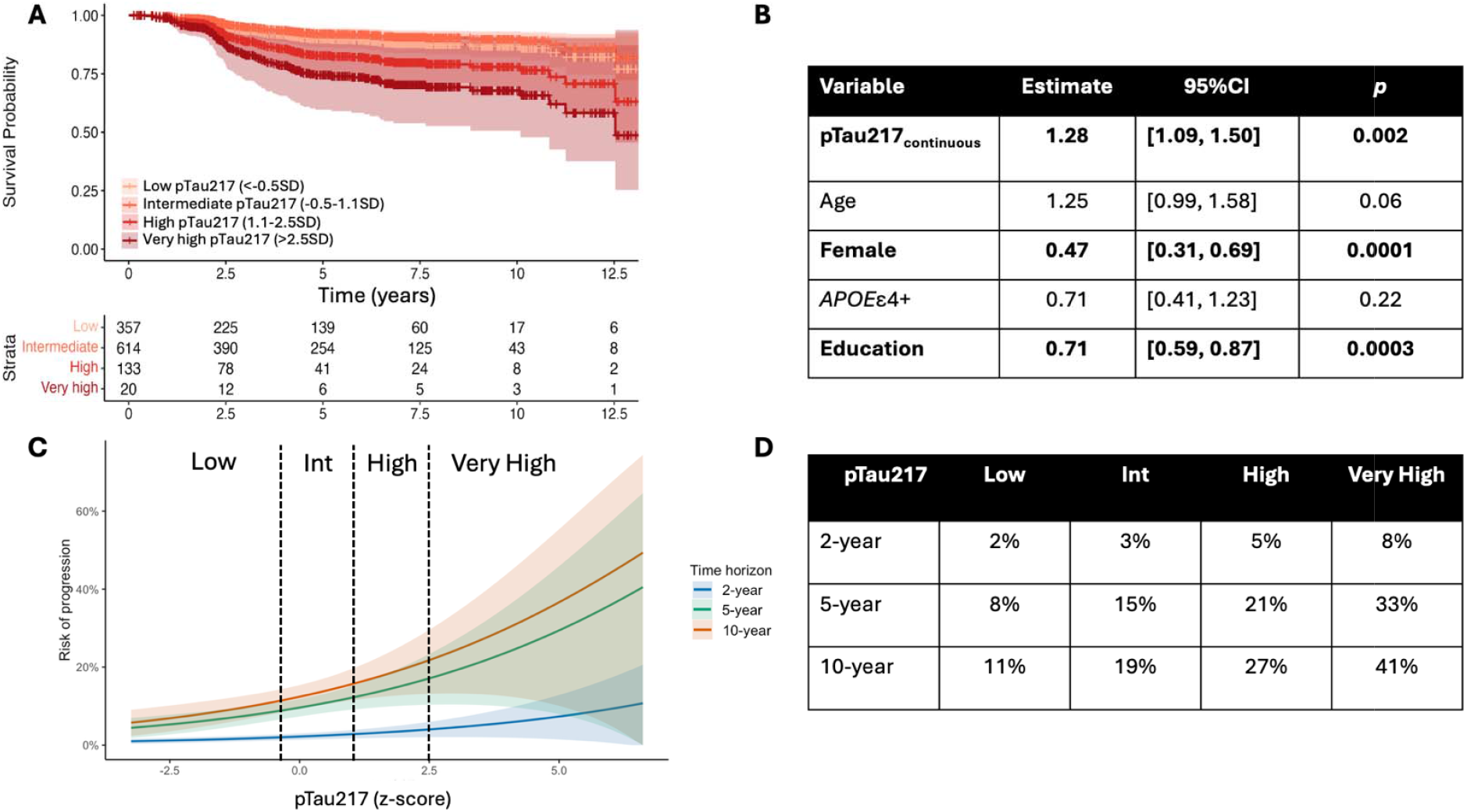
In <10 AβCL only (n=1,124, events=111): (A) Survival curves for clinical progression to cognitive impairment by p-tau_217_. (B) Standardized model estimates of the main effects of predictors of interest in the survival model (all continuous predictors were standardized [mean = 0, SD = 1]). (C) Model-based 2-, 5-, and 10-year absolute risk by p-tau_217_ category (Low (corresponding to <10 AβCL), Intermediate (10-24 AβCL), High (25-60 AβCL) and Very High (>60 AβCL). (D) Tabular absolute risk values by p-tau217 group

### Association between p-tau217, Aβ-CL, and subsequent decline on longitudinal cognitive testing

Longitudinal associations were estimated using generalized least squares models with natural cubic splines for time, incorporating an autoregressive correlation matrix to account for within-participant correlation across time. These models examined the association between continuous and categorical p-tau217 and cognitive decline on the harmonized latent Preclinical Alzheimer Cognitive Composite (lPACC). Our primary model examined p-tau217 and covariates (age, sex, education, *APOE*_ε_4 status) over time in the model. A subsequent model examined the additional impact of Aβ-CL over time. Due to power constraints, we ran our models only in the full sample. Full model estimates can be found in Supplementary Table 5A-C.

In spline-based longitudinal models (n=22,309 observations), baseline plasma p-tau217 significantly modified lPACC trajectories over time (likelihood ratio [LR] against a model without p-tau217*spline time terms: χ^2^ = 278.4, *p*<0.0001; see Figure 4A), indicating faster cognitive decline with increasing p-tau217 levels. By contrast, p-tau217 was not associated with baseline lPACC, suggesting its primary influence is on longitudinal change. The rate of decline was greatest at higher levels of p-tau during early follow-up (see Figure 4B). Individuals with low p-tau217 (–0.5SD) showed an annualized increase of +0.023 lPACC/year [SE=0.003], whereas those with intermediate p-tau217 (+1.1SD) showed an annualized decline of 0.01 lPACC/year [SE=0.01]. Both the High and Very-High p-tau217 groups (+2.5SD and +6SD, respectively) showed a much more marked annualized decline of –0.036 [SE=0.0045] and –0.104 lPACC/year [SE=0.01], respectively (all p < 10 ^−^□). The spline explained 22.9% of the variance in longitudinal lPACC. Compared with a covariates-only model model, inclusion of baseline plasma p-tau217 significantly improved model fit (LR test: χ^2^ = 505.8, p < 0.0001). The incremental explanatory contribution of p-tau217 was further quantified using a LR-based partial R^2^, indicating that p-tau217 explained approximately 2.3% of outcome variation beyond the covariates-only model.

**Figure 4.**
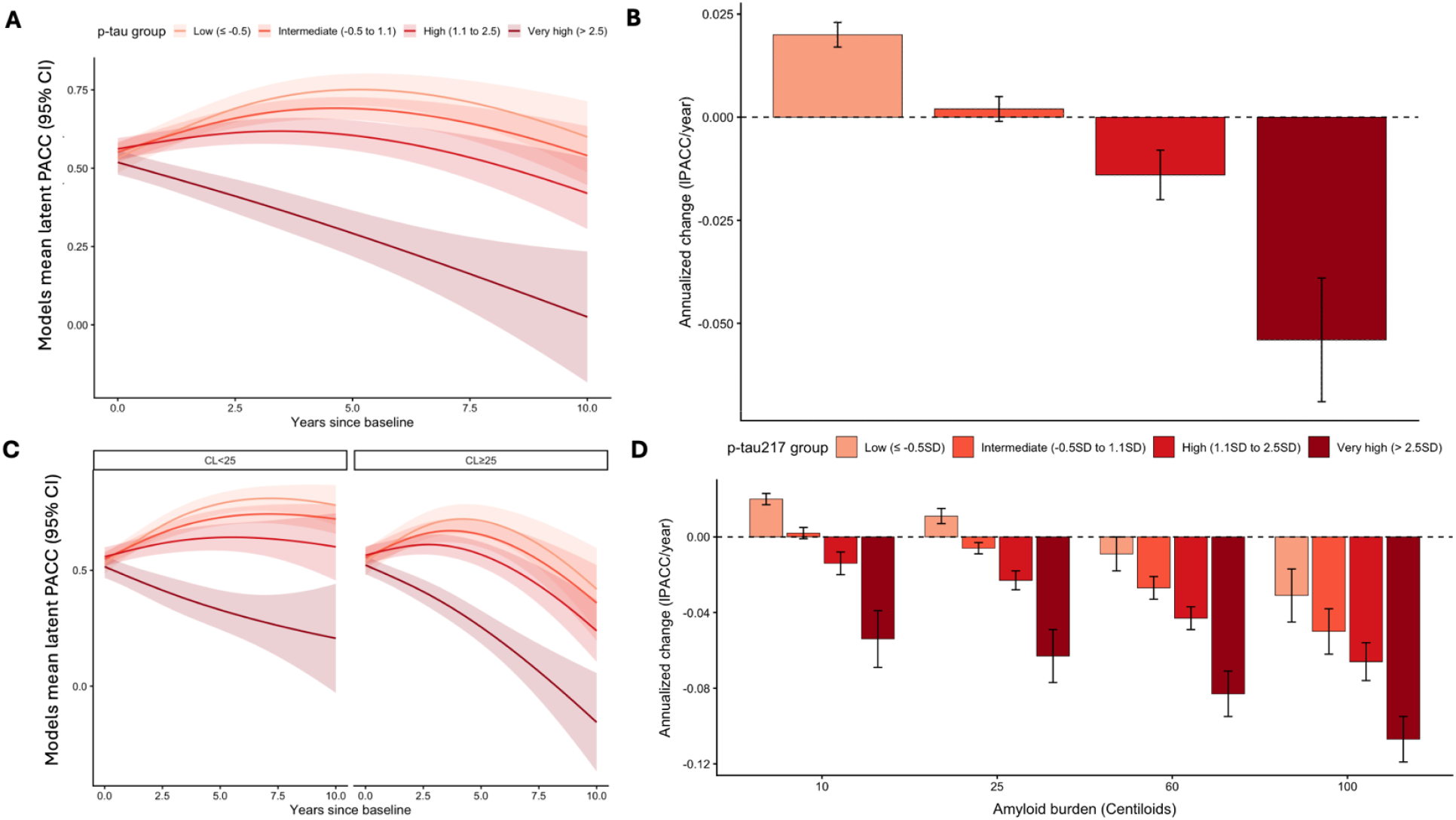
Baseline plasma p-tau217 and Aβ CL independently modify longitudinal cognitive decline. (A) Modeled mean lPACC trajectories over 10 years by baseline plasma p-tau217 group (Low,Intermediate, High, Very High). Shaded bands represent 95% confidence intervals. (B) Estimated average annualized change in lPACC by p-tau217 group derived from spline models. (C) Modeled trajectories stratified jointly by p-tau217 group and Aβ status (Centiloid <25 vs ≥25). (D) Estimated average annualized decline across the p-tau217 continuum at representative Aβ CL (10, 25, 60, and 100 CL).

When modeling both baseline plasma p-tau217 and Aβ-CL with spline terms over time, both variables independently modified longitudinal lPACC trajectories (LR against a model without spline interaction terms: χ^2^ = 290.6, *p*<0.0001; see Figure 4C). Higher p-tau217 levels were associated with progressively faster decline across all Aβ-CL levels, while lower p-tau217 levels were associated with a practice effect that became attenuated over time. The model demonstrated a marginal R^2^ of 23.0%, and the AIC was significantly lower, suggesting an improvement when including Centiloid in the model (log-likelihood ratio: 12.82, *p*=0.005). At a much more severe level of Aβ-CL burden (100 CL), individuals in the highest p-tau217 group declined at −0.11 lPACC/year, compared to −0.03 lPACC/year in the lowest p-tau217 group. P-tau217 was found to drive decline even at 10 Aβ CL, with Very High p-tau217 levels associated with annualized decline of -0.055 lPACC/year (see Figure 4D). We also examined a model that interacted p-tau217 and Aβ-CL over time. In this model, we found significant interaction terms indicating that higher Aβ-CL and higher p-tau217 was associated with faster rates of lPACC decline.

## Discussion

In this large, multi-cohort study of more than 2,700 cognitively unimpaired older adults, we show that plasma p-tau217 provides robust prognostic information across clinically relevant time periods. Higher baseline p-tau217 levels were associated with substantially increased risk of progression to cognitive impairment, with individuals in the very high group (p-tau217 > 2.5SD corresponding to >60 Aβ-CL) reaching an estimated 10-year risk of approximately 61% [95% CI: 53-68%]. Absolute risk over 2 years remained low across the p-tau217 continuum, even in individuals with markedly elevated p-tau217 values, underscoring the prolonged preclinical phase of AD. By contrast, striking risk gradients emerged over 5 and 10 years, which is unsurprising given the insidious nature of the clinicopathological trajectory of AD. This temporal dissociation underscores the popular notion of a critical window for intervention, during which pathological processes are already active but have not yet translated into overt cognitive impairment.

P-tau217 remained independently associated with risk of progression after adjusting for major risk factors for dementia: age, sex, Aβ-CL and *APOE*ε4 status. While Aβ-PET and *APOE*ε4 are well-established predictors, our results indicate that plasma p-tau217 reflects both complementary and unique pathological processes that meaningfully contribute to prognosis^20, 21^. The high AUCs and C-indices remained largely unchanged with the inclusion of Aβ-CL, supporting that AUC is relatively insensitive once the performance of the model is already high. Given that model fit was markedly better with the inclusion of Aβ-CL values, this should not be interpreted as lack of biological relevance or clinical importance.

We specifically examined individuals with low Aβ burden (<10 Aβ-CL) to evaluate whether plasma p-tau217 retains prognostic value in the apparent ‘absence’ of substantial fibrillar amyloid pathology. This analysis addresses a clinically relevant question, as blood-based biomarkers are increasingly used in individuals who have not undergone Aβ-PET imaging, and for whom Aβ status will be unknown. Demonstrating whether p-tau217 predicts progression even in the setting of very low Aβ-CL informs the potential utility of p-tau217 as an early and independent risk marker^22^, rather than simply a proxy for established Aβ-CL burden. Among this group, associations between p-tau217 and progression were attenuated and confined largely to the extreme upper tail of the p-tau217 distribution (i.e., discordant cases). These findings must be interpreted cautiously, however, due to fewer progression events and wider confidence intervals at this end of the continuum, but they raise important biological questions. Elevated p-tau217 in individuals with very low Aβ-PET may reflect several, non-mutually exclusive explanations, including early pathological processes (secretion of p-tau217 in response to early Aβ plaque deposition), assay variability, or atypical or non-AD mechanisms that merit further investigation. This may also arise from methodological limitations in PET quantification at the lowest Centiloid ranges. For instance, signal-to-noise characteristics differ across tracers, particularly among some ^18^F compounds where white-matter binding is a well-characterized phenomenon and can obscure cortical signal at the lower end of the Aβ-PET distribution^23-26^ relative to ^11^C-PiB/^18^F-NAV4694. Thus, some individuals classified as <10 CL, particularly from the ^18^F tracers, may harbor biologically meaningful fibrillar Aβ that does not yet exceed conventional PET positivity thresholds. Further, it is also possible that the limit of detection in Aβ-PET is simply higher than for p-tau217 suggesting that signal from peripheral circulating p-tau might be indicating the presence of fibrillar Aβ that does not yet meet the traditional PET positivity threshold^27^. As a result, discordance between plasma p-tau217 and Aβ-PET in this range may reflect measurement error on either side of the biomarker spectrum rather than true pathological dissociation. Studies suggest that Aβ-PET signal below traditional thresholds (e.g., <20 CL) confer important predictive information about future cognitive decline^16^, supporting the notion that even lower levels of Aβ-PET correspond to clinically-meaningful risk. Substantially greater sampling in this discordant range, from studies such as the Alzheimer’s Plasma Extension Study (APEX)^22^, will be essential to clarify the prognostic and biological significance of these edge cases.

These data have several practical implications. First, they reinforce that graded levels of pathological burden, rather than binary positive/negative classifications, convey substantially more prognostic information about the timing of clinical progression, as has been shown previously for Aβ-PET across the Centiloid spectrum^13^. Second, plasma p-tau217 represents as a scalable blood-based biomarker which can provide prognostic information beyond Aβ-PET, enabling refined risk stratification and individualized risk communication for CU older adults undergoing biomarker testing. Third, framing risk in absolute terms over defined time horizons can help contextualize biomarker abnormalities for CU patients. Absolute risk estimates can also guide monitoring strategies and identifying individuals who may benefit from closer follow-up. Individuals with very high p-tau217 levels and elevated long-term risk may benefit from repeat biomarker testing or earlier referral to specialty care, whereas those with low predicted risk may be reassured and monitored less intensively. Fourth, the strong long-term prognostic performance supports the use of p-tau217 currently being used for trial enrichment, allowing investigators to optimize screening thresholds to maximize event rates while minimizing screen failures. Similar approaches have already been successfully implemented in recent prevention trials, such as AHEAD 3-45^1^. More broadly, these findings reinforce the emerging role of blood-based biomarkers as scalable tools for pre-symptomatic risk stratification, bridging the gap between research and future clinical practice. As multiplex proteomic assays become increasingly available, plasma-based platforms may ultimately support multivariable risk models that integrate complementary biological pathways and proteomic candidates, further advancing precision medicine approaches in AD.

Our findings build directly on prior efforts to quantify absolute risk of cognitive impairment using biomarker information to estimate lifetime risk of AD dementia in cognitively unimpaired individuals using NIA-AA preclinical staging criteria^11^, and a more recent publication by Jack and colleagues^13^, seeking to model continuous Aβ-PET severity (using Centiloid values) and estimating both lifetime and 10-year absolute risk of MCI and dementia. The former demonstrated differences in risk by age and biomarker-defined stage and found that 10-year absolute risk of dementia remained relatively low for most preclinical states, but that baseline age exerted a strong modifying effect^13^. Unlike the current study, the authors classified dementia using epidemiologic questionnaire criteria, which typically correspond to moderate to late-stage dementia, whereas our focus was on the earlier disease continuum. Notably, they reported an absolute 10-year risk of approximately 45% to MCI in a 75-year-old with very elevated Aβ-PET (100 Aβ-CL). This estimate was attenuated relative to the risk we observed (61% [95% CI: 53-68%] among individuals with very high p-tau217 levels corresponding to >60 Aβ-CL). It is possible that p-tau217 is representing signal associated with the severity of underlying Aβ and tau pathology (i.e., Aβ bothering the tau) rather than Aβ status *per se*. Finally, a recent study from 2,148 older adults recruited from a community-based cohort in Sweden^14^, showed that the top two quartiles of p-tau217 were associated with significantly higher hazard ratios than the bottom reference quartile, however, they did not provide absolute risk estimates. They did report high NPVs and low PPVs across all the serum analytes examined, supporting the high prognostic utility of p-tau217 to predict risk of progression.

Beyond clinical diagnostic progression, very high p-tau217 levels were also associated with faster longitudinal cognitive decline, even after accounting for Aβ-PET levels. Individuals with very high p-tau217 may represent a biologically distinct disease stage beyond just reflecting Aβ-PET burden alone, consistent with AT framework models in which different biological processes drive risk across the disease continuum. We found our strongest effects in those with Very High p-Tau217, corresponding to an estimated >60 AβCL, which has is associated with the earliest evidence of tau-PET proliferation beyond the medial temporal lobe^19^. Given our use of natural cubic splines to accommodate non-linear cognitive trajectories, direct quantitative comparison with studies assuming linear decline is not straightforward, however, previous studies have reported consistent associations between plasma p-tau217 and rates of global cognitive decline^28, 29^, including with the PACC^30, 31^. Other studies assuming non-linearity have reported similar findings^31^. Together, the concordance between our diagnostic and cognitive outcomes further supports p-tau217 as a sensitive marker of disease processes in preclinical stages.

Across all models, male sex emerged as a consistent predictor of progression to clinical impairment and faster rates of cognitive decline. Although this finding may appear counterintuitive given the higher lifetime prevalence of AD among women, some prior studies suggest that women demonstrate greater cognitive resilience in preclinical stages^32, 33^, and yet other studies have reported no sex difference in incidence rates below the age of 90^34-36^. Importantly, sex was modeled here as a fixed effect rather than in interaction with other risk factors, and we therefore cannot infer sex-specific vulnerability to diagnostic outcome via p-tau217. Much of our work on female-specific vulnerability to tau and rates of cognitive decline focuses primarily on those who already demonstrate elevated Aβ-PET burden^37-40^. Ongoing and future work will explicitly examine interactions between sex, age, *APOE*ε4, p-tau217, and amyloid burden to determine whether the observed sex-specific risk is reflected by differential profiles of resilience across the disease continuum.

Several limitations warrant consideration. Clinically actionable thresholds for p-tau217 among CU individuals remain to be established, and although we anchored cut points to biologically meaningful Aβ-CL ranges, further work is needed to define values that would clearly alter clinical management or trial eligibility. Relatedly, our analyses were conducted across multiple assay platforms and values were harmonized using standardization. While this approach enabled pooling across cohorts and improved statistical power, it also introduces inherent limitations, and our results should not be interpreted as providing platform-specific clinical cut points. Future work will be needed to establish assay-specific thresholds and evaluate their clinical utility in independent samples. Our models did not explicitly incorporate comorbidities, vascular pathology, or competing risks, although we adjusted for major demographic and genetic risk factors and leveraged large, well-characterized cohorts with standardized longitudinal follow-up, which increases confidence in the robustness of the observed associations. Unlike prior work that explicitly modeled out-of-study events and mortality, our analyses relied on in-study outcomes, raising the possibility of informative censoring at longer time horizons, whereby individuals who discontinue follow-up may differ systematically in their underlying risk of progression compared with those who remain under observation. The general consistency of findings across cohorts and other sensitivity analyses supports the overall stability of our risk estimates. It is important to note, however, that only a small fraction of the sample (5%) was followed longer than 10 years, which can result in reduced reliability of our estimates at that end of the time scale. Further, these longer follow-up intervals were primarily contributed to by a subset of cohorts with extended observation periods, and we therefore interpret these 10-year estimates in the context of cohort-specific analyses that demonstrated broadly consistent effects. One exception was the WRAP study, which included particularly extended follow-up; however, this cohort was younger at baseline and experienced relatively few events, limiting its influence on long-term estimates. Although we had representation of more diverse cohorts in this study, racial and ethnic diversity remains a constraint on generalizability, highlighting the need for validation in more representative samples, particularly given known disparities in dementia risk and access to biomarker testing. Indeed, the inclusion of these observational and prevention trial cohorts with distinct recruitment and eligibility criteria may introduce a degree of selection bias, potentially limiting the generalizability of our estimates. Finally, future work should focus on refining multivariable prediction models that integrate additional biomarkers and clinical features.

In summary, plasma p-tau217 provides robust long-term prognostic information for individuals who are currently cognitively unimpaired and lays the groundwork for future development of individualized risk prediction similar to lipid and blood pressure levels used for cardiovascular risk scores. By translating levels of biomarker abnormality into absolute risk estimates, this work moves the field closer to clinically meaningful pre-symptomatic risk stratification, supporting both trial design, and ultimately shared decision making in the clinic should disease modifying therapies become available for this population.

## Methods

### Participants

We pooled data from multiple well-characterized observational and prevention cohorts that enrolled cognitively unimpaired older adults with baseline plasma p-tau217 and PET imaging and longitudinal clinical follow-up. Data from 2,705 cognitively unimpaired participants (mean age 69.8 years [SD 7.0; range 46.0–98.1], 1,704 women [63%]) were harmonized across six cohorts: the Alzheimer’s Disease Neuroimaging Initiative (ADNI), the Anti-Amyloid Treatment in Asymptomatic Alzheimer’s (A4) Study and the adjacent observational the Longitudinal Evaluation of Amyloid Risk and Neurodegeneration (LEARN) Study, the Harvard Aging Brain Study (HABS), the Wisconsin Registry for Alzheimer’s Prevention (WRAP), and the Health & Aging Brain Study-Health Disparities (HABS-HD; see Supplementary Table 6 for public data access links). Cohort-specific demographic and clinical characteristics are summarized in Table 1. Across cohorts, participants were predominantly female and highly educated, with a pooled mean of 16.0 years of education (SD 3.2). *APOE*ε4 carriership was present in 39% of the sample, with substantial variability across cohorts, reflecting differences in recruitment strategies. Overall, 43% of participants were Aβ+ at baseline, with marked enrichment in A4 and lower prevalence in LEARN, HABS-HD and WRAP, consistent with cohort design. Racial and ethnic composition varied substantially across cohorts, with greater representation of Black/African American and Hispanic participants in HABS-HD, while other cohorts were predominantly White. We defined our primary outcome of interest as progression to mild cognitive impairment (MCI) or dementia, or the presence of two consecutive CDR global scores of 0.5 in cohorts where formal diagnoses were unavailable. Event rates varied across cohorts, reflecting differences in follow-up duration, age distribution, and recruitment criteria.

All contributing studies were approved by their respective institutional review boards, and the present analyses were conducted under the ethical guidelines stipulated by the Massachusetts General Brigham Institutional Review Board. All participants provided written informed consent.

### Plasma p-tau217

Methods and assays for plasma p-tau217 quantification within each cohort are summarized in Supplementary Table 7. The following immunoassay platforms were used: the Alzpath assay on the Quanterix HD-X platform (WRAP)^41^, the Fujirebio Lumipulse and Janssen R&D Simoa platform (ADNI), the Meso Scale Diagnostics (MSD) S-Plex assay on the QuickPlex SQ120 (A4/LEARN and HABS). To harmonize plasma p-tau217 levels across cohorts, we implemented a previously published method to log10-transform and standardize values within each cohort using z-scores derived within each cohort from Aβ-participants aged <70 years^4^. Extreme outliers (<Q1 − 5×IQR or >Q3 + 5×IQR) were excluded prior to standardization to facilitate harmonization across cohorts. We treated p-tau217 as a continuous predictor as well as a categorical predictor. To do this, we anchored the grouping thresholds to meaningful Aβ-CL thresholds of 10, 25 and 60 Aβ-CLs using a conditional quantiling approach. Specifically, we identified the distribution of p-tau217 values conditional on each Aβ-CL PET threshold and selected p-tau217 cut points corresponding to those Centiloid-defined risk strata. This strategy allows plasma p-tau217 categories to be calibrated against well-characterized Aβ-PET severity levels, rather than relying on arbitrary percentile-based thresholds. In brief, this approach identifies the p-tau217 values that correspond to specific levels of Aβ burden by estimating the distribution of p-tau217 conditional on each CL threshold. It is particularly useful in a multi-cohort, multi-platform setting, where absolute p-tau217 values vary across assays and populations. By conditioning on Aβ-PET burden, we ensured that p-tau217 categories reflected comparable biological risk states across cohorts, improving interpretability and external validity. The resulting p-tau217 thresholds were -0.5SD, 1.1SD and 2.5SD, corresponding to Low (n=594), Intermediate (n=1,089), High (n=594) and Very High (n=506). By using these cut-offs, we found a very strong alignment between p-tau and CL groups (χ^2^(9)=1354.2, *p*<0.0001; see Supplementary Table 4A). A demographics table comparing these groups can be found in Supplementary Table 2.

### Aβ Positron Emission Tomography

Each cohort used different PET tracers: ADNI, A4/LEARN and HABS-HD used ^18^F-Florbetapir, while ADNI also used ^18^F-Florbetaben, and HABS and WRAP used ^11^C-Pittsburgh Compound B (PiB). The PET acquisition parameters for each study have been published previously^42-46^, and further details can be found in Supplementary Table 8. For the purposes of this study, we converted all within-cohort standardized uptake value ratios (SUVr) and distribution volume ratios (DVR from the logan graphical method) to Centiloid using within-cohort established conversion algorithms. For all cohorts, we considered the baseline Aβ-PET scan which was closest (limited to within 2 years) to each individual’s first blood plasma p-tau217 data.

### Clinical diagnosis

The primary outcome was incident cognitive impairment, defined as a within-cohort diagnosis of mild cognitive impairment (MCI) or dementia. Across cohorts, diagnostic determination was based on established study-specific procedures, which in most cases involved multidisciplinary clinical consensus panels integrating cognitive testing, functional assessments, and clinical judgment. In cohorts where formal adjudicated diagnoses were unavailable or inconsistently recorded, progression was operationalized as the occurrence of two or more consecutive Global Clinical Dementia Rating (CDR) scores ≥0.5, consistent with emerging functional impairment. Although diagnostic procedures varied slightly across studies, all cohorts employed annual follow-up for longitudinal clinical assessment, and diagnoses were made blinded to plasma biomarker status.

### Harmonization of Preclinical Alzheimer Cognitive Composite (PACC) variants

A table of all tests used to harmonize into a latent Preclinical Alzheimer Cognitive Composite can be found in Supplementary Table 9. We used different variants of the Preclinical Alzheimer’s Cognitive Composite (PACC)^47^ as the cognitive outcome. Below, we discuss in detail our harmonization method, which we have previously published to demonstrate the robust approach that we take to ensure equivalence. In HABS, the PACC was constructed from five tests: Free Recall plus Total (sum of free and cued) score from the Free and Cued Selective Reminding Test (FCSRT), delayed paragraph recall from Logical Memory IIA, the Digit-Symbol Substitution Test (DSST) from the Wechsler Adult Intelligence Scale-Revised, the Mini-Mental Status Examination (MMSE) and Categories (Animals, Fruits and Vegetables)^47^. A4/LEARN, however, does not include Categories, and so the standard four-test version of the PACC was examined^48^. In WRAP, the PACC was created from three cognitive tests; it used the total score across trials of the Rey Auditory-Verbal Learning Test (AVLT), and Logical Memory II and Digit Symbol Substitution Test^49^. For ADNI, a five-test version of the PACC was created from ADAS-Cog Word Recall, Logical Memory IIA, Trails B, and the MMSE^48^. Finally, HABS-HD possessed a five-test version of the PACC, using the Spanish-English Verbal Learning Test (SEVLT), which has been shown to have very good correspondence with English-only verbal learning tests. HABS-HD also included Logical Memory IIA, DSST, Categories (Animals) and MMSE^50^.

The harmonization method for the PACC has been published in detail^51-54^. In brief, we harmonized PACC scores across cohorts using an item response theory-based approach implemented through confirmatory factor analysis, in which all neuropsychological tests were modeled as indicators of a common latent cognitive factor. Tests shared across cohorts (e.g., MMSE, Logical Memory Delayed Recall) were treated as anchors, with their parameters constrained to be equal across studies, thereby linking cohorts to a common metric. Item-level data were used where available, allowing differential weighting of test components and avoiding the strong assumptions inherent to simple z-score standardization. This approach yields directly comparable cognitive scores across cohorts while preserving meaningful between-study differences in baseline performance and longitudinal change

### Statistical Analyses

Analyses were performed using R (version 4.5.0). We used Cox proportional hazards models to examine the association between plasma p-tau217 and risk of progression to cognitive impairment. Time-to-event was defined as time from baseline assessment to first occurrence of cognitive impairment, with censoring at last available follow-up. Death was not available in these cohorts and so was not included in our models. Primary models were fitted in the full sample and included plasma p-tau217 as either continuous or categorical (Low, Intermediate, High, Very High), adjusting for cohort, age, sex, years of education, and *APOE*ε4 status. Subsequent models included baseline Aβ-PET Centiloid (Aβ-CL) values to assess the independent contribution of plasma p-tau217 beyond Aβ-PET burden. Secondary analyses were conducted in participants with very low Aβ burden (<10 Aβ-CL) using the same modeling framework. For each Cox model, we estimated absolute risk of progression at 2, 5, and 10 years using model-based survival predictions for a reference individual defined by the sample-average covariate profile. Model discrimination and calibration were evaluated using 5-fold cross-validation, including time-dependent area under the curve (AUC) at 10 years, Harrell’s concordance index (C-index), and Brier score.

To characterize longitudinal cognitive trajectories, we used generalized least square models incorporating natural cubic splines for time^55^, consistent with prior methodological work. This approach allows flexible modeling of non-linear change over time while preserving interpretability and statistical efficiency. Baseline plasma p-tau217 was modeled in interaction with spline-expanded time terms (df=2) to estimate differential rates and acceleration of lPACC change. All models were adjusted for the same covariates as above. Additional models incorporated baseline Aβ-CL values and tested p-tau217 × Aβ-CL × spline-expanded time term interactions to evaluate synergistic effects on cognitive trajectories. The number and placement of spline knots were selected *a priori* following published recommendations^56^ and confirmed using Akaike Information Criterion. Model fit was summarized using marginal R^2^ for generalized least squares models, reflecting variance explained by fixed effects. Effect estimates are reported with standard errors.

## Data Availability

All data produced are available online at:
A4/LEARN https://www.synapse.org/Synapse:syn61250768/wiki/628717
ADNI https://ida.loni.usc.edu/login.jsp
HABS https://www.synapse.org/Synapse:syn53910452/wiki/626438
HABS-HD https://apps.unthsc.edu/itr/reports
WRAP https://wrap.wisc.edu/data-requests-2/

https://www.synapse.org/Synapse:syn61250768/wiki/628717

https://ida.loni.usc.edu/login.jsp

https://www.synapse.org/Synapse:syn53910452/wiki/626438

https://apps.unthsc.edu/itr/reports

https://wrap.wisc.edu/data-requests-2

