## Supplementary Table 1 for "The prognostic value of blood-based p-tau217 levels on progression to clinical impairment"

**Supplementary Figure A.** Wave diagram of follow-up, with censoring events colored in purple in (A) the full sample, and (B) each cohort.

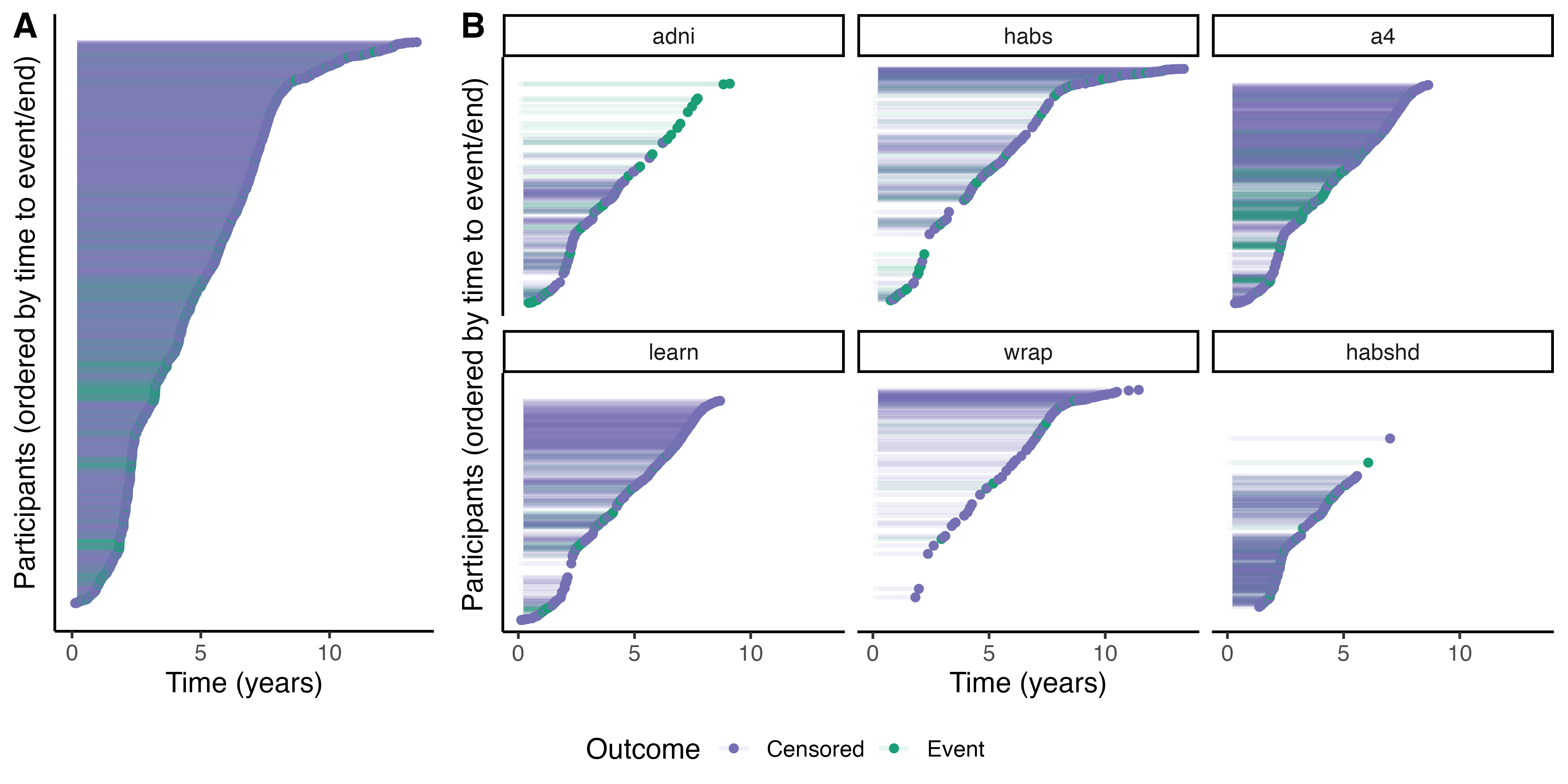

**Supplementary Table 1.** Breakdown of progression to first event categories (i.e., time to event to MCI, dementia, 2+ >=0.5CDR, or remaining stable at the end of the study) by cohort

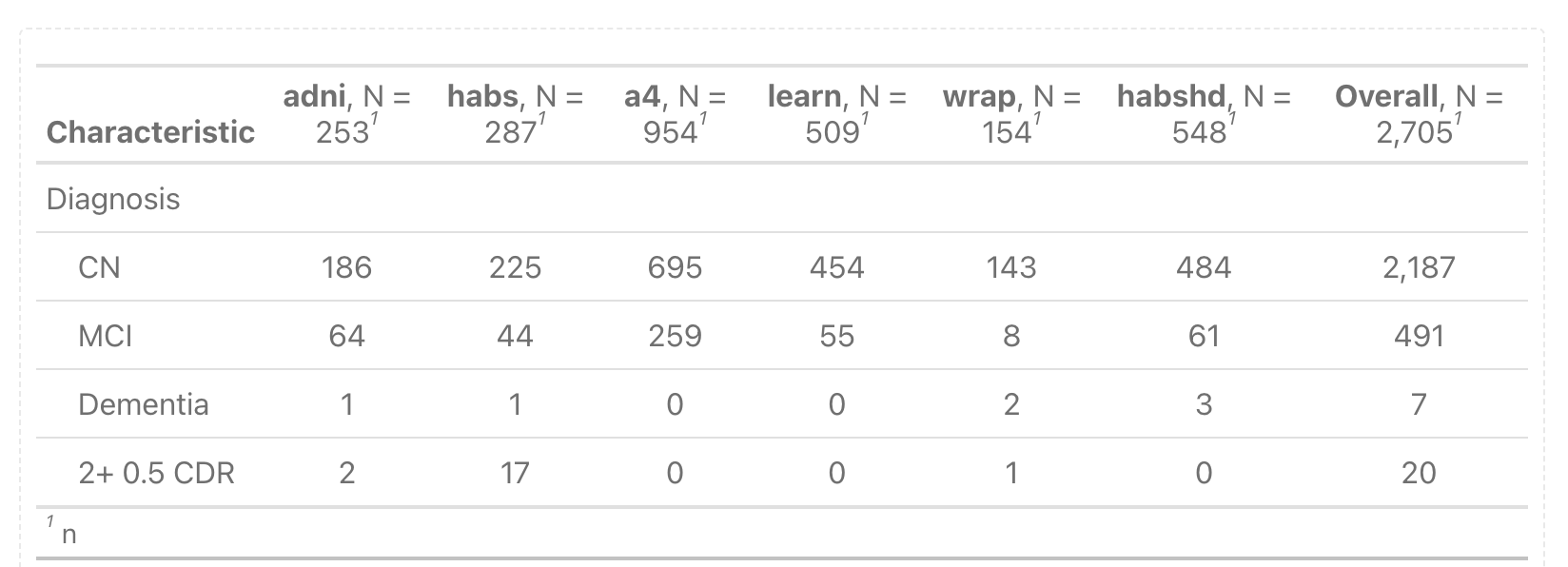

**Supplementary Figure B.** Construction and characterization of plasma p-tau217 groups. (A) Boxplot distribution of plasma p-tau217 levels across amyloid burden categories defined by Centiloid thresholds (<10, 10–25, 26–60, >60). (B) Global p-tau217 quartile thresholds applied across Centiloid stages. Points represent individual participants, colored by p-tau217 group (Low, Intermediate, High, Very High). Dashed horizontal lines indicate global quartile cut-points used to define p-tau217 groups. (C) Continuous relationship between Aβ-CL burden and plasma p-tau217. Grey points represent individual observations. The solid blue line shows the linear fit, while the dark red curve represents a non-linear (loess) fit with 95% confidence interval (shaded). Vertical dashed lines indicate Centiloid thresholds used for amyloid staging. Horizontal red lines indicate global p-tau217 quartile cut-points. (D) Kernel density curves show the distribution of z-scored plasma p-tau217 levels stratified by a ‘traditional’ Aβ status (A− vs A+) cutpoint defined using 25 CL. Vertical dashed lines indicate the pre-specified p-tau217 cut-points (−0.47, 1.13, and 2.53) used to define Low, Intermediate, High, and Very High p-tau217 groups.

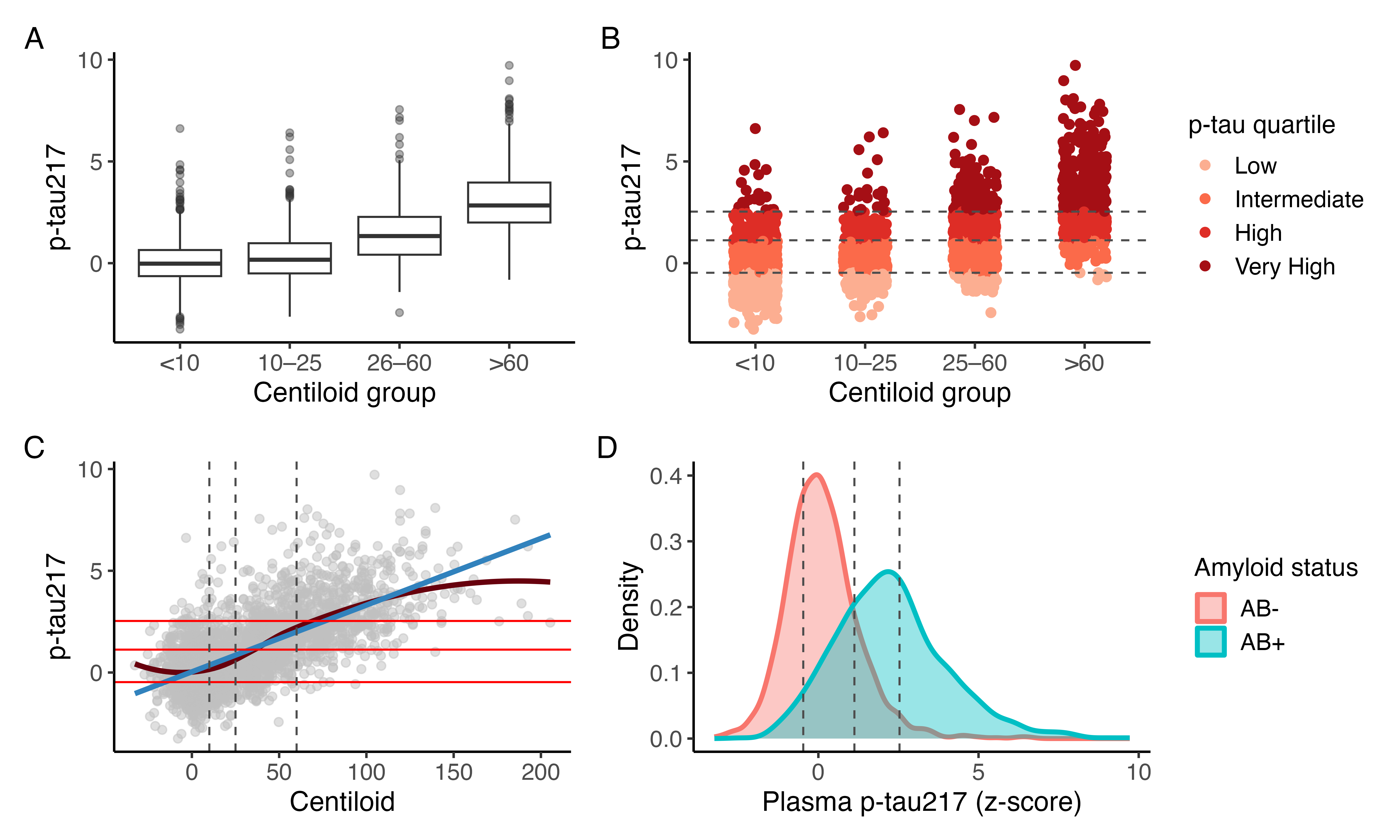

**Supplementary Table 2**. Sample characteristics by p-tau217 group

| **Characteristic** | **Overall**  N = 2,705^1^ | **Low (<10CL cut-off)**  N = 516^1^ | **Intermediate (10–25CL cut-off)**  N = 1,089^1^ | **High (26–60CL cut-off)**  N = 598^1^ | **Very High (>60CL cut-off)**  N = 502^1^ | **p-value**^2^ |
| --- | --- | --- | --- | --- | --- | --- |
| **Age** | 69.80 (7.01) [46.00, 98.08] | 66.81 (7.39) [46.00, 88.25] | 68.57 (6.86) [50.00, 98.08] | 71.65 (6.26) [49.00, 90.14] | 73.35 (5.55) [52.00, 90.41] | <0.001 |
| **Sex** |  |  |  |  |  | 0.007 |
| Female | 1,706 (63%) | 353 (68%) | 695 (64%) | 352 (59%) | 306 (61%) |  |
| **Education** | 16.01 (3.20) [0.00, 30.00] | 15.60 (3.44) [0.00, 30.00] | 16.03 (3.22) [0.00, 26.00] | 16.05 (3.06) [2.00, 25.00] | 16.32 (3.02) [0.00, 27.00] | 0.007 |
| **APOE** |  |  |  |  |  | <0.001 |
| e4+ | 1,059 (39%) | 112 (22%) | 332 (30%) | 306 (51%) | 309 (62%) |  |
| **Aβ Status** |  |  |  |  |  | <0.001 |
| Aβ+ | 1,157 (43%) | 43 (8.3%) | 247 (23%) | 405 (68%) | 462 (92%) |  |
| **p-tau217** | 1.04 (1.77) [-3.25, 9.72] | -1.03 (0.49) [-3.25, -0.47] | 0.30 (0.45) [-0.47, 1.12] | 1.78 (0.41) [1.13, 2.53] | 3.89 (1.23) [2.54, 9.72] | <0.001 |
| **Follow-up** | 5.19 (2.73) [0.12, 13.50] | 5.07 (2.78) [0.13, 13.50] | 5.16 (2.85) [0.12, 12.94] | 5.15 (2.64) [0.53, 13.06] | 5.43 (2.50) [0.32, 13.21] | 0.070 |
| **Progression events** | 518 (19%) | 47 (9.1%) | 115 (11%) | 140 (23%) | 216 (43%) | <0.001 |

**Supplementary Table 3.** Full model estimates for the Cox Proportional Hazard Ratios predicting progression to cognitive impairment: (A) p-tau217 + covariates, (B) p-tau217 + Aβ-CL, (C) in <10 Aβ-CL only: p-tau217 + covariates

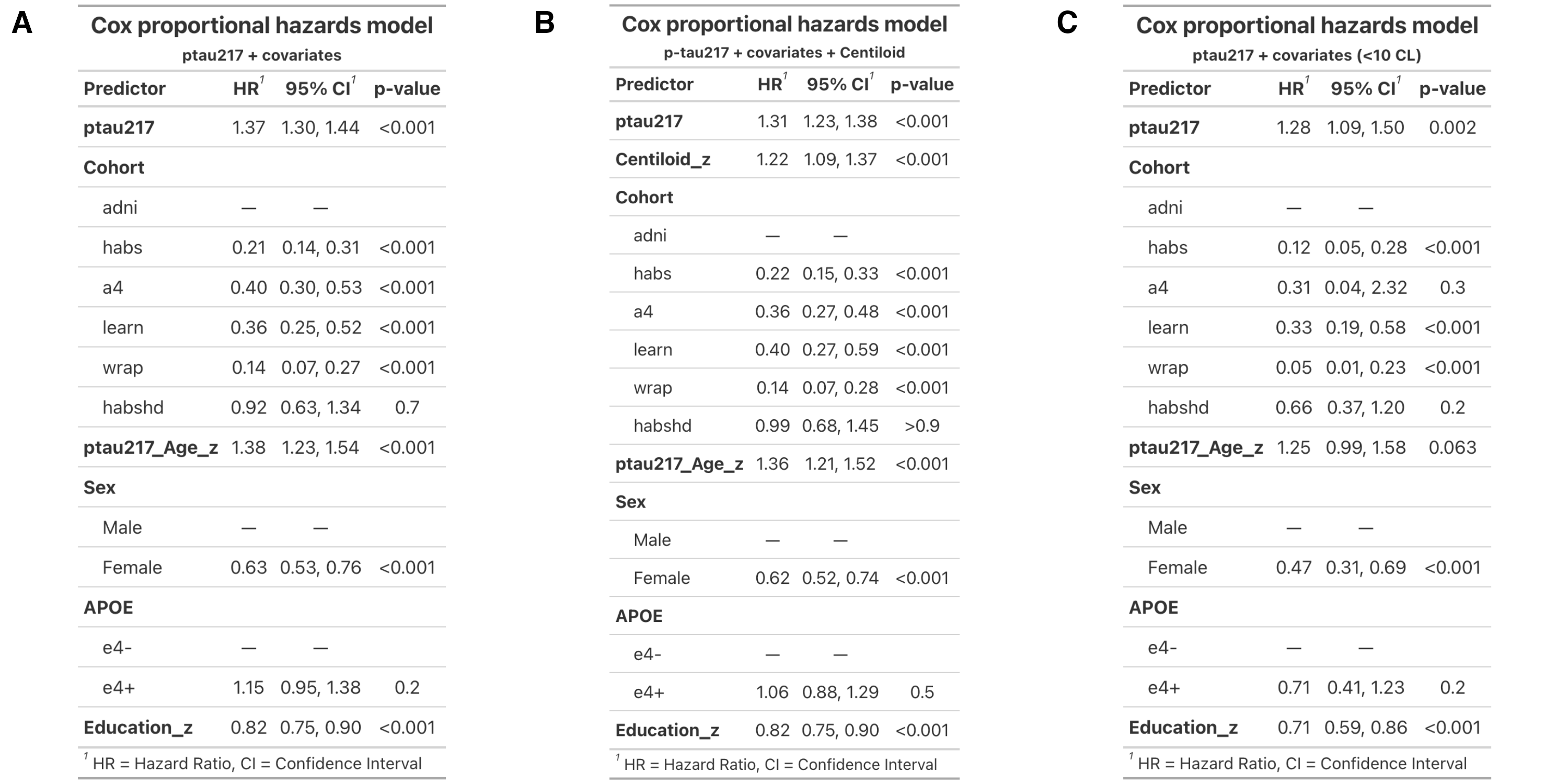

**Supplementary Figure C.** Forest plot showing cohort-specific hazard ratios (HRs) and 95% confidence intervals for [ptau217]. Points represent HRs from Cox proportional hazards models fitted within each cohort. The pooled estimate (triangle) was obtained using fixed-effects meta-analysis. The vertical dashed line denotes HR=1. All cohorts demonstrate consistent directionality of effect, with the pooled estimate indicating a significant association.

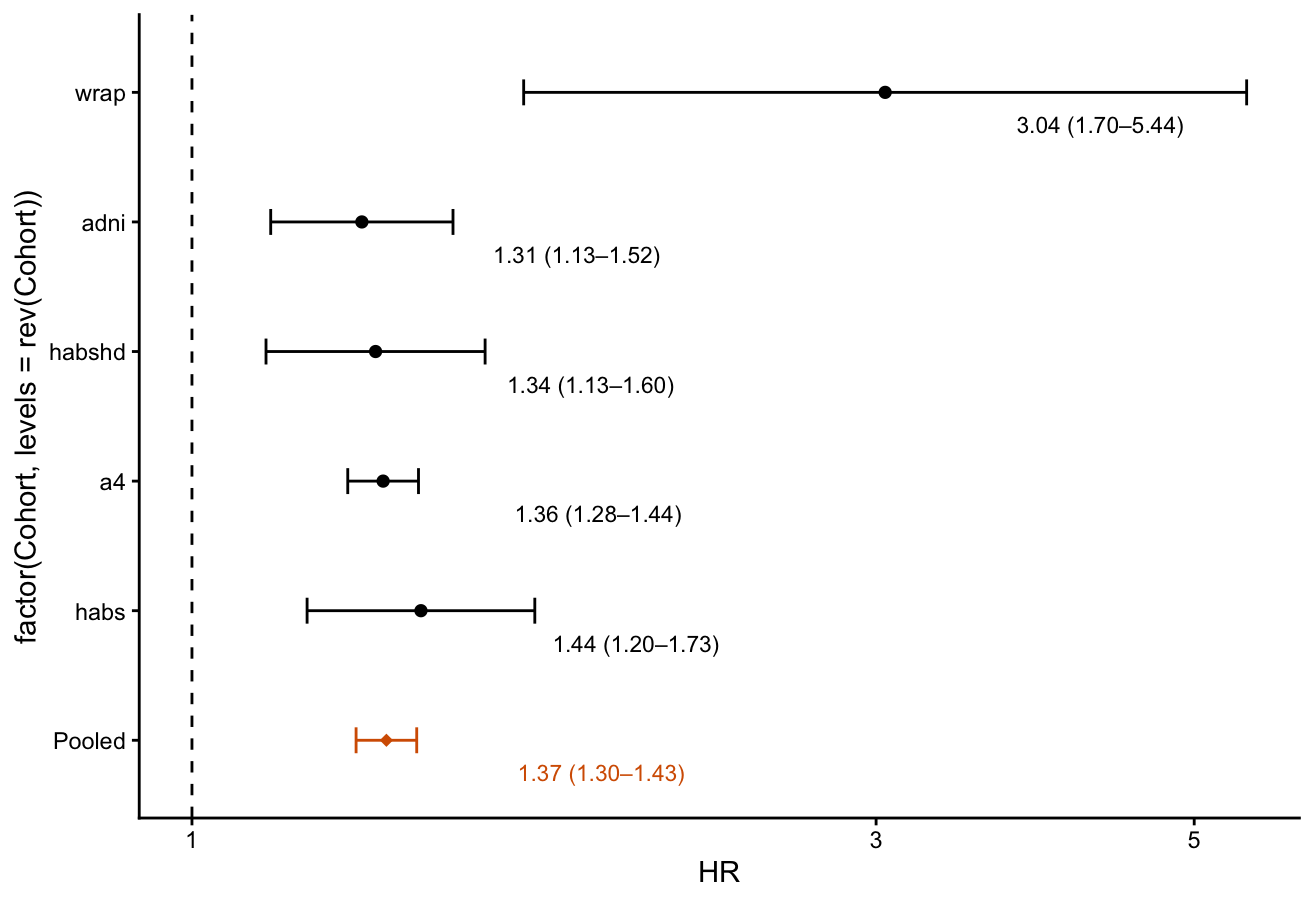

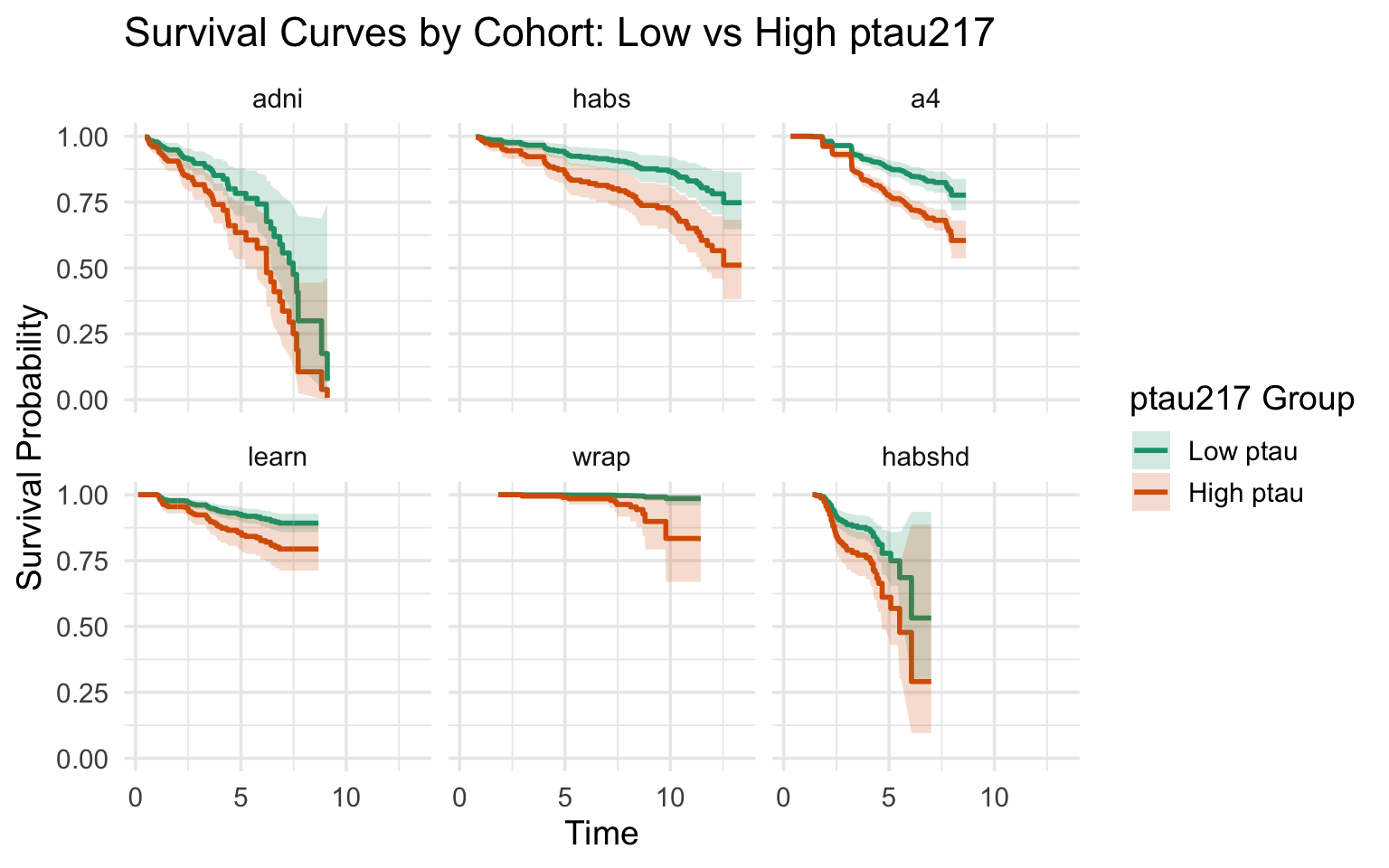

p-tau217 group

-1SD

+1SD

**Supplementary Table 3.** Cohort-specific Cox Proportional Hazard Ratio models

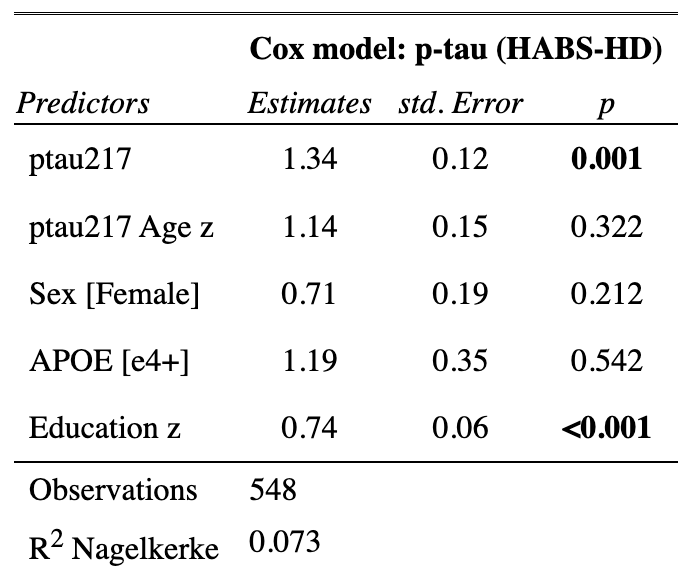

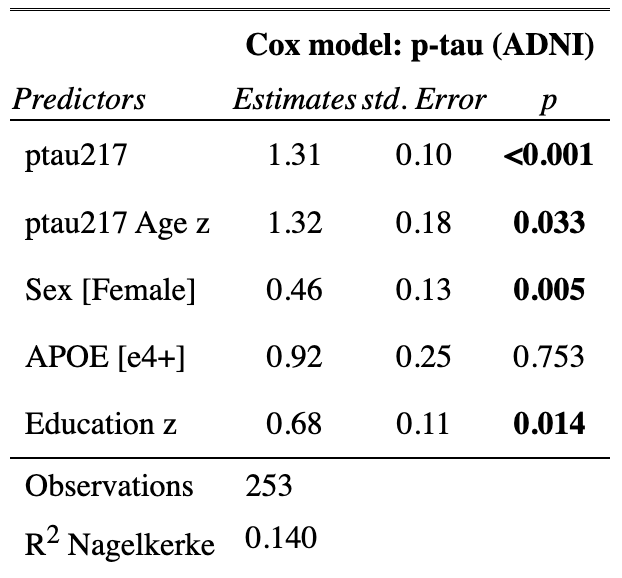

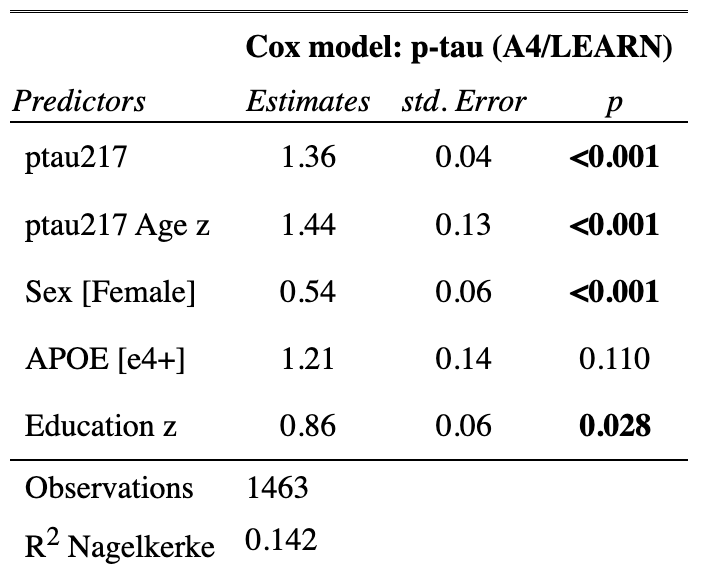

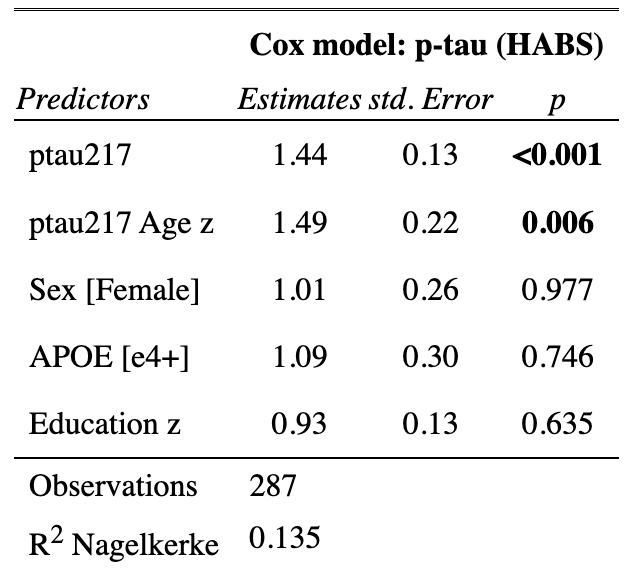

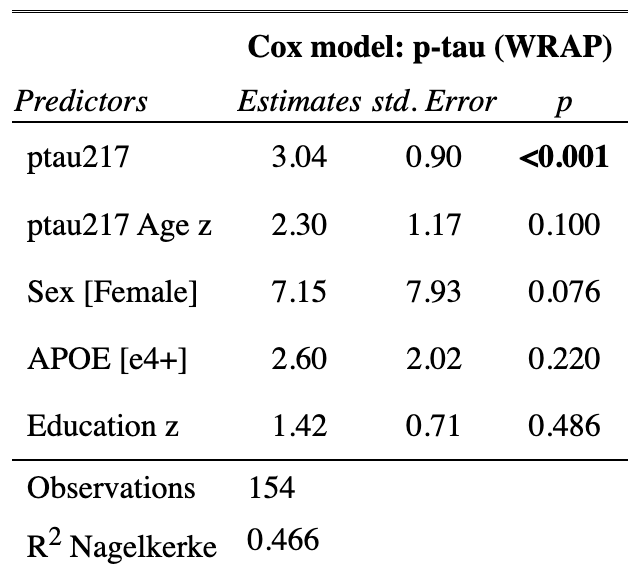

**Supplementary Table 4.** Contingency table between p-tau217 groups, CL groups and events in (A) full sample and (B) <10 Aβ-CL

**(A)**

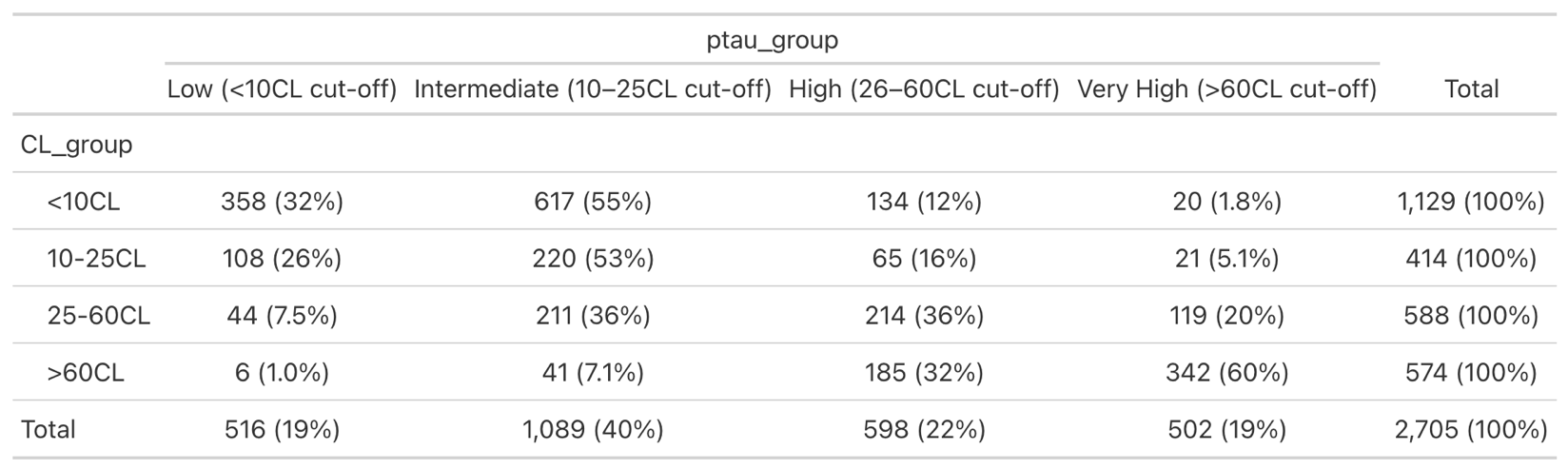

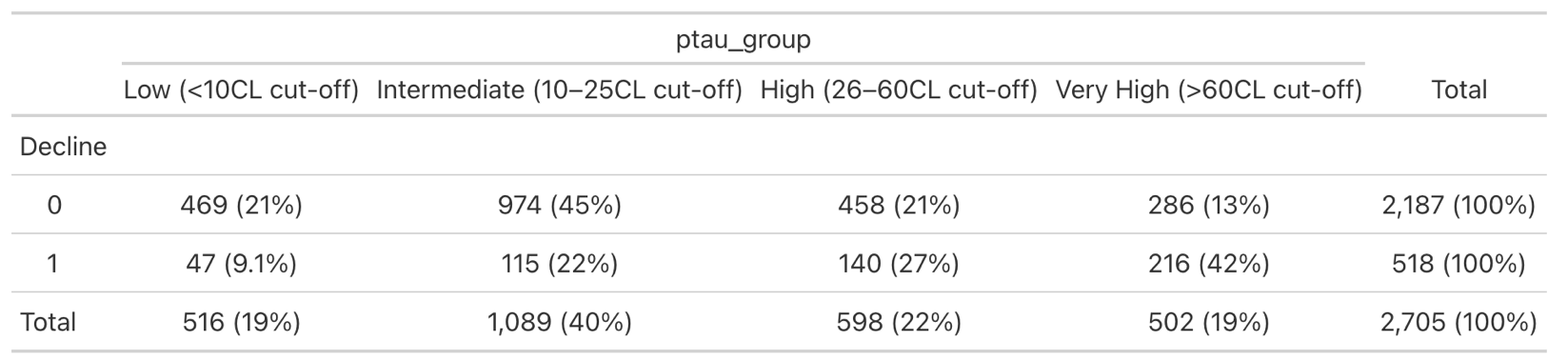

Note: Decline: 0 denotes no progression to cognitive impairment within the study, 1 denotes progression to cognitive impairment within the study

**(B)**

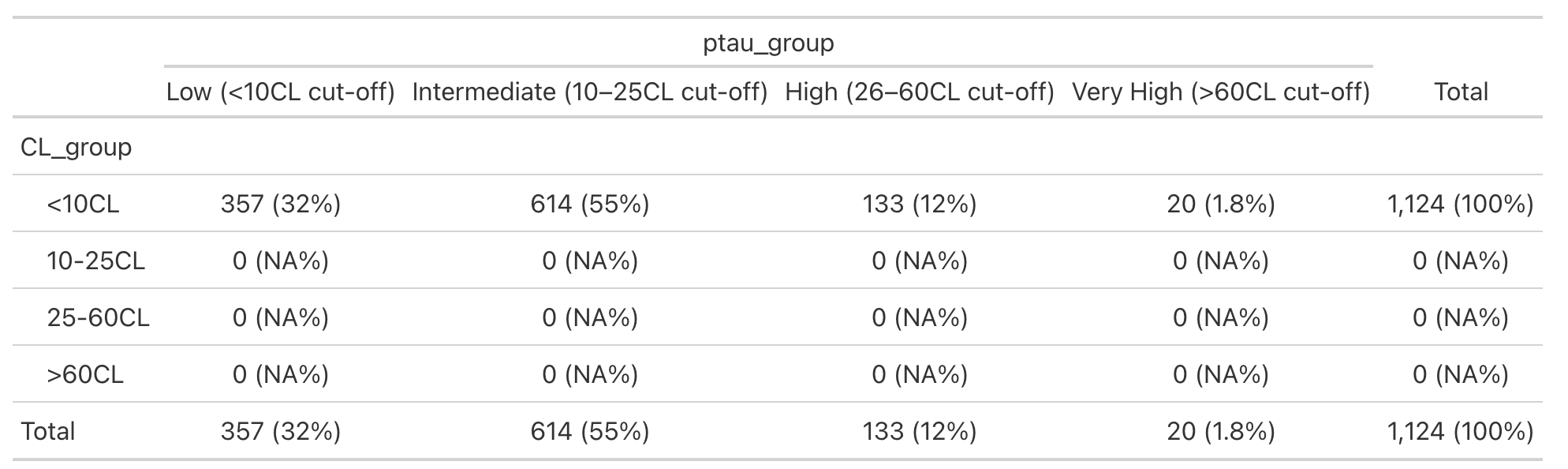

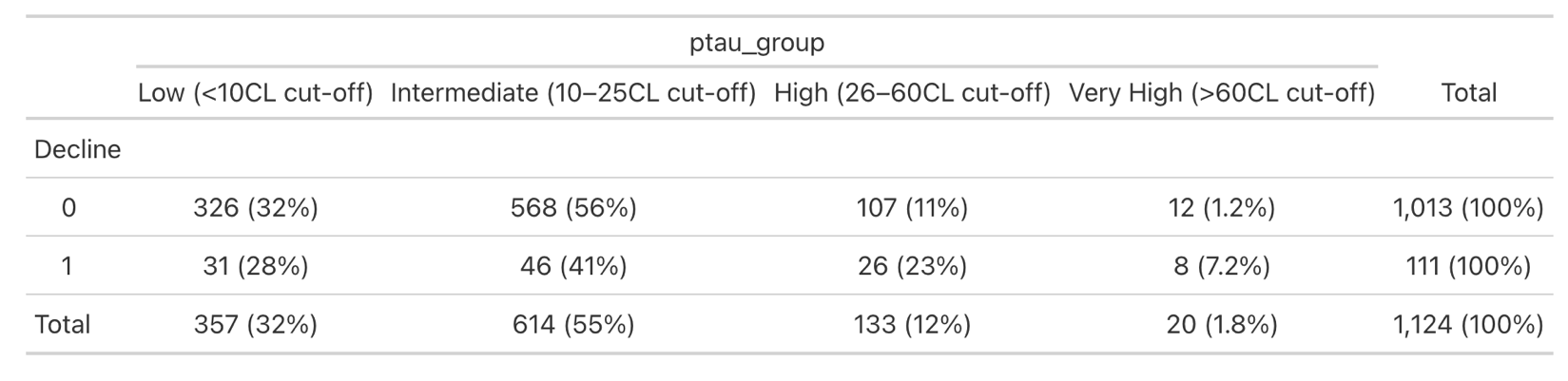

Note: Decline: 0 denotes no progression to cognitive impairment within the study, 1 denotes progression to cognitive impairment within the study

**Supplementary Table 5.** Natural Cubic Spline models with full estimated terms for (A) p-tau217 only, (B) p-tau217 and Centiloid, and (C) the interaction between p-tau217 and Centiloid

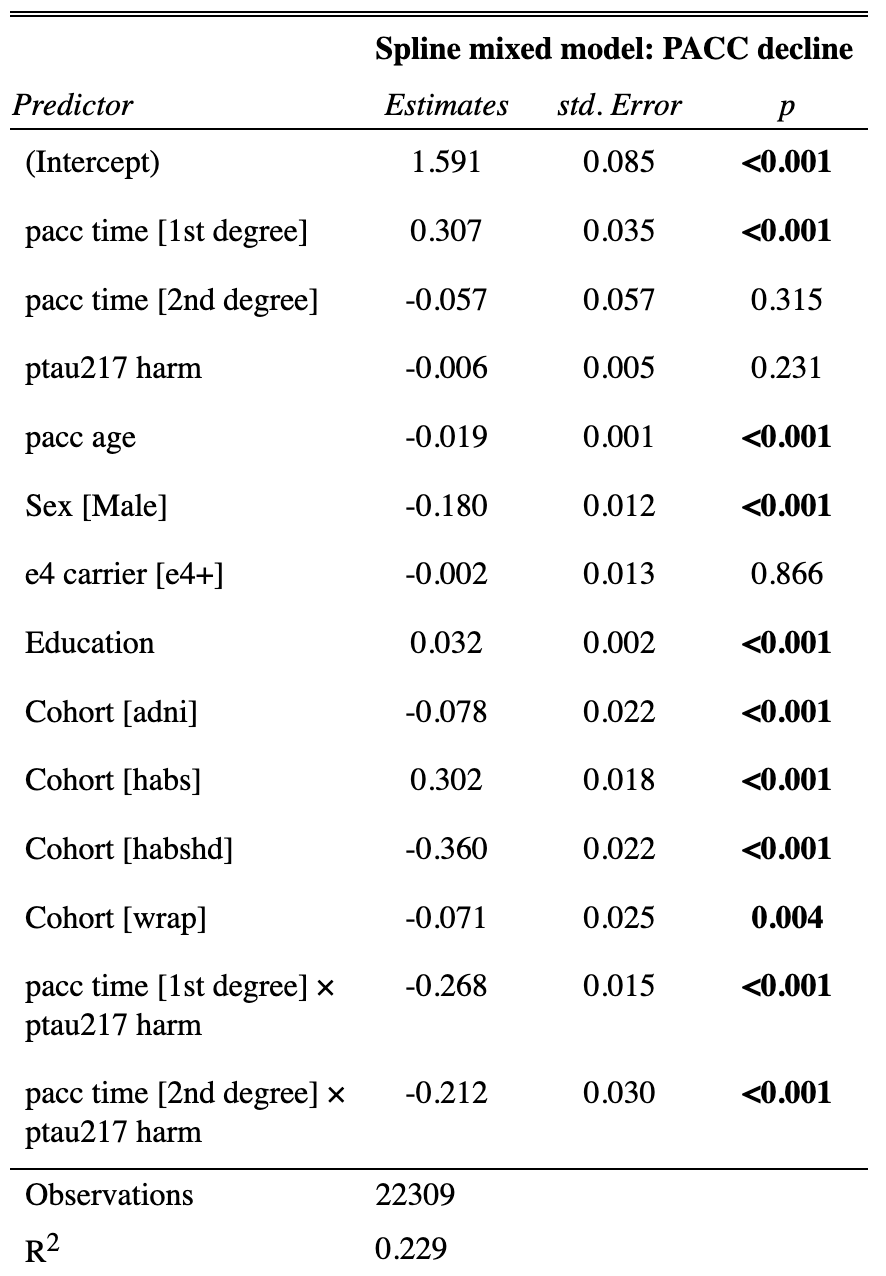

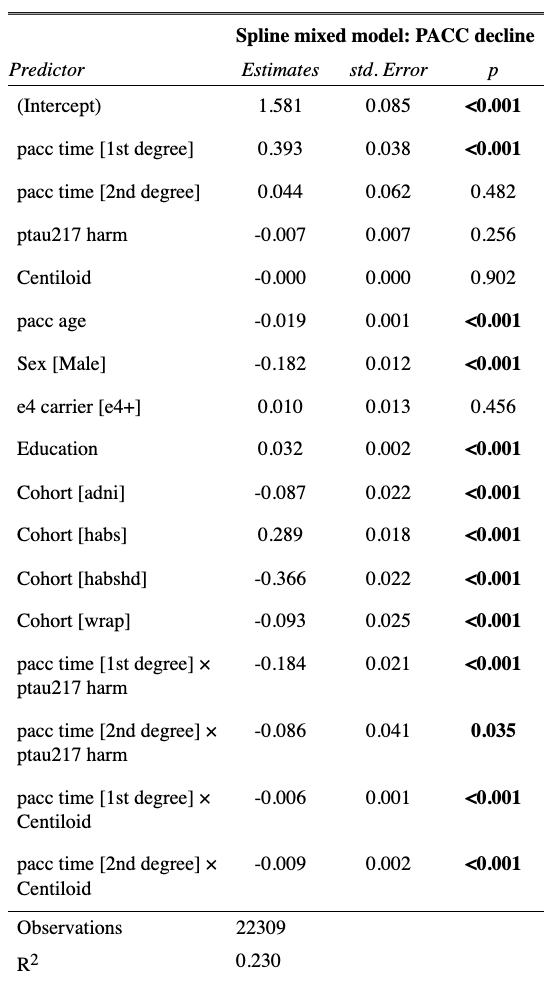

**B**

**A**

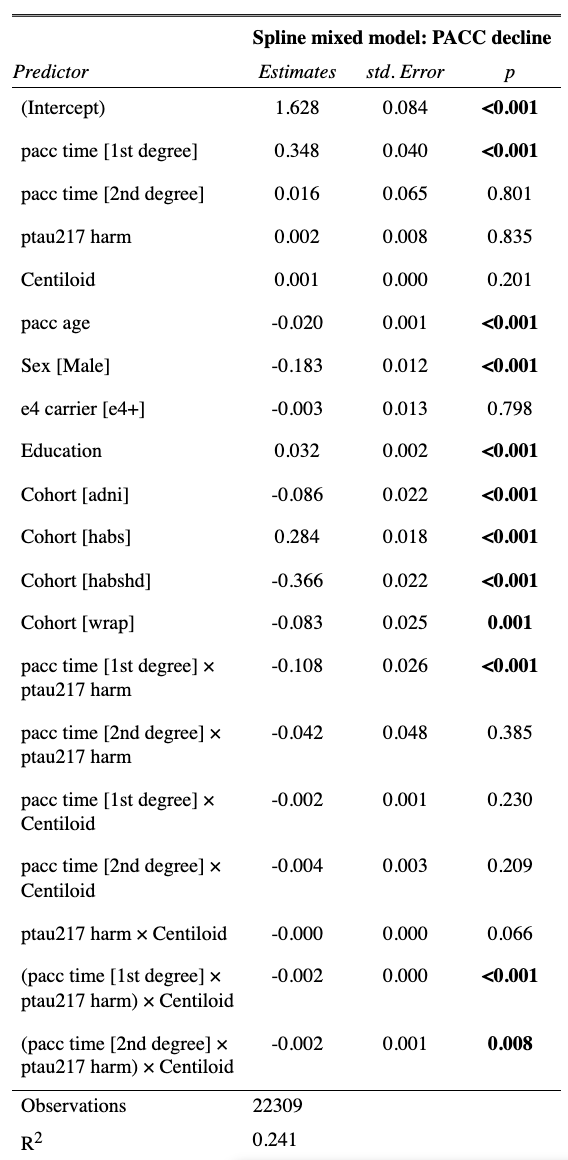

**C**

**Supplementary Table 6.** Public data access links for each cohort

| **Cohort** | **Data Access Link** |
| --- | --- |
| A4/LEARN | https://www.synapse.org/Synapse:syn61250768/wiki/628717 |
| ADNI | https://ida.loni.usc.edu/login.jsp |
| HABS | https://www.synapse.org/Synapse:syn53910452/wiki/626438 |
| HABS-HD | https://apps.unthsc.edu/itr/reports |
| WRAP | https://wrap.wisc.edu/data-requests-2/ |

**Supplementary Table 7.** Ptau217 collection and assay methods by cohort

| **Cohort** | **Ptau217 Assay** | **Collection methods** |
| --- | --- | --- |
| A4/LEARN | MSD | Quantification of P-tau217 was assayed on an analytically validated ECL immunoassay using an MesoScale (MSD) at the CAP-accredited, CLIA-certified Lilly Clinical Diagnostics Laboratory on plasma samples.^1^ |
| ADNI | UPENN:  Fujirebio Lumipulse G1200 automated immunoassay platform  Janssen:  Quanterix Simoa pTau217 v2 (ALZpath) assay on the Simoa HD-X Automated Immunoassay Analyzer | UPENN: All blood samples were collected in EDTA collection tubes, processed to produce plasma, transferred to a transfer tube as described in the ADNI4 Procedures manual V2.0. The 0.5 mL aliquots prepared from each sample were stored at -80^0^C until the day of analysis. On the day of analysis, each thawed (one-half hour at room temperature).  Samples were sent from UPenn to Quanterix (Billerica, MA), acting as subcontractor for Janssen R&D Plasma. Plasma p-tau217 concentrations were quantified using the commercial ALZpath p-Tau217 immunoassay on the Single-Molecule Array (SIMOA) HD-X instrument. |
| HABS | MSD S-Plex assay on the QuickPlex SQ120 reader | Overnight fasted blood draw. Plasma samples were collected into dipotassium ethylenediaminetetraacetic acid (K2EDTA) tubes, centrifuged, and frozen within 2 h of collection, and the aliquots of plasma were stored at −80°C until use. Plasma samples sent to the second laboratory (MIND Biomarker Core at Mass General Brigham) were analyzed using ultrasensitive S-PLEX assay kits (MSD, Meso Scale Diagnostics, Rockville, MD, USA) to quantify p-tau217 (CAT# K151APFS), employing a sandwich immunoassay format using monoclonal antibodies and electrochemiluminescence detection, following previously published protocols^2, 3^. |
| HABS-HD | Quanterix Simoa HD-X Automated Immunoassay Analyzer | Released with v7 (r7_) dataset. Samples were assayed in the University of North Texas Health Science Center Institute for Translational Research (ITR) Laboratory by the ITR Biomarker Core. Fasting blood samples are collected as part of the HABS-HD protocol and include the collection of plasma (using EDTA acid tubes). Collection and processing are completed per international pre-analytic guidelines and include a 2 hour stick-to-freezer timeline.^4^ Stored in a biorepository (−80° freezer) until processing. Proteomic assays for this study were processed on a multiplex biomarker assay platform using electrochemiluminescence (ECL) using commercially available kits from Quanterix. |
| WRAP | Simoa pTau217 v2 (ALZpath) assay on the Quanterix HD-X Automated Immunoassay Analyzer | Thirty millilitres of blood was drawn from each participant into 310 mL lavender top EDTA tubes (BD 366643; Franklin Lakes, New Jersey, USA). Samples were mixed gently by inverting 10–12 times and were centrifuged 15 min at 2000 g at room temperature within 1 h of collection. Plasma samples were aliquoted into 2 mL cryovials (Wheaton Cryoelite W985863; Millville, New Jersey, USA). Aliquoted plasma was frozen at −80°C within 90 min and stored for up to 10 years. |

**Supplementary Table 8.** Aβ-PET Scanner (Where Applicable) and Acquisition Methods

| **Cohort** | **Amyloid PET Tracer** | **PET Scanner** | **Acquisition protocol** |
| --- | --- | --- | --- |
| A4/LEARN | [18F] florbetapir | N/A (multi-site) | Acquired 50 to 70 minutes after receiving an injection of 10 mCi of florbetapir F 18 and measured using a mean cortical standardized uptake value ratio (SUVr) with a whole cerebellar reference region. |
| ADNI | [18F] florbetaben, [18F] florbetapir, and [18F] NAV4694 | N/A (multi-site) | Florbetapir images con-  sisted of 4 x 5 min frames acquired at 50–70 min after injection, which  were realigned, averaged, resliced to a common voxel size (1.5 mm3), and  smoothed to a common resolution of 8 mm3 in full width at half maximum. SUVrs referenced to whole cerebellum.  For Florbetaben,  a 20-min PET scan (4 × 5 min frames) starting at least 90 min after intravenous injection of 300 MBq ± 20% of 18 F-Florbetaben. SUVrs referenced to cerebellar gray. |
| HABS | Pittsburgh compound B (PiB) | Siemens/CTI ECAT HR Scanner | After injection of 8.5-15 mCi 11C PiB, 60-minutes of dynamic data were acquired in 3D acquisition mode and reconstructed in 39 frames (8 x 15s, 4 x 60s, and 27 x 120s). Distribution volume ratios (DVRs) referenced to cerebellar gray. |
| HABS-HD | [18F] florbetaben | MCT20 (Siemens MCT 20 PET/CT) or Vision (Siemens BioGraph Vision 450 PET/CT). | Study partners are injected with an 8.1 mCi (±10%) bolus of florbetaben. A 20 minute (4 frames of 5-min each) dynamic emission acquisition is started 90 min post-injection following the acquisition of a low-dose CT scan used for attenuation correction. SUVrs referenced to cerebellar gray. |
| WRAP | Pittsburgh compound B (PiB) | Siemens EXACT HR+ | Amyloid imaging is conducted with [C-11] Pittsburgh Compound-B (PiB) PET using a dynamic 70-minute protocol. DVRs referenced to cerebellar gray. |

**Supplementary Table 9.** Cognitive test batteries administered and used to form the harmonized Preclinical Alzheimer’s Cognitive Composite (PACC). The table summarizes domain-specific neuropsychological measures used to assess global cognition, memory, verbal fluency, list learning, and executive function within each cohort. Purple denotes the overlapping tests that were able to be anchored across cohorts using item response theory. This item response theory has been published previously.

|  | **ADNI** | **HABS** | **A4/LEARN** | **WRAP** | **HABS-HD** |
| --- | --- | --- | --- | --- | --- |
| **Global** | Total MMSE Score | Total MMSE Score | Total MMSE Score |  | Total MMSE Score |
| **Memory** | Logical Memory Delayed Recall (Anna Thompson Story) | Logical Memory Delayed Recall (Anna Thompson Story) | Logical Memory Delayed Recall (Robert Miller Story*) | Logical Memory Delayed Recall (Anna Thompson Story) | Logical Memory Delayed Recall (Anna Thompson Story) |
| **Verbal fluency** | Category fluency – Animals | Category fluency – Animals +Vegetables + Fruits |  |  | Category fluency – Animals |
| **List-Learning** | ADAS-Cog Del Word Recall | Free and Cued Selective Reminding Test (FCSRT) | Free and Cued Selective Reminding Test (FCSRT) | Rey Auditory-Verbal Learning Test (AVLT) | Spanish-English Verbal Learning Test (SEVLT) |
| **Executive function** | Trails B score | Digit Symbol Substitution Test (WAIS-R) (90sec) | Digit Symbol Substitution Test (WAIS-R) (90sec) | Digit Symbol Substitution Test (WAIS-R) (90sec) | Digit Symbol Substitution Test (WAIS-R) (90sec) |
